# Generation and Evaluation of Realistic Synthetic Clinical Progress Notes for Prostate Cancer using Large Language Models

**DOI:** 10.64898/2026.05.25.26354027

**Authors:** Álvaro Rey-Blanes, Francisco J. Moreno-Barea, Javier Veredas-Morente, Eloy Vivas-Vargas, Fátima Gil-García, Francisco J. Veredas

## Abstract

**Background and Objective:** Access to real-world electronic health records (EHRs) remains limited by privacy, governance and annotation constraints, hindering the development of clinical natural language processing models. Realistic synthetic progress notes may provide EHR-like corpora that preserve clinically rigorous information on diagnoses, treatments, symptoms, imaging, laboratory findings and therapeutic trajectories without relying directly on sensitive patient records. This study evaluates whether large language models (LLMs) can generate realistic Spanish prostate cancer progress notes from published case reports, preserving clinical content, temporality and hospital-style conventions.

**Methods:** We compiled 109 Spanish prostate cancer case reports from the biomedical literature and characterised their clinical content using Spanish biomedical named-entity recognition (NER) models, complemented by rule-based extraction of prostate specific antigen (PSA) values and Gleason scores. GPT-5.4 Nano, Qwen 3.5:35B A3B and GLM-5 were used to generate EHR-style progress notes from these case reports under plain-text and entity-enriched prompting strategies, in both zero-shot and few-shot settings. Evaluation combined lexical and semantic similarity metrics with structured LLM-as-a-judge assessment using Claude Sonnet 4.6, binary safety screening and expert clinical review.

**Results:** All models preserved substantial clinical content, although lexical-overlap metrics showed variable agreement with semantic and clinical quality assessments, reflecting the abstractive nature of the task. Entity-enriched prompting improved lexical and semantic alignment, but did not consistently improve clinical safety. Qwen 3.5:35B A3B was unstable under entity-enriched few-shot prompting, showing increased safety-critical errors and contradictions. GPT-5.4 Nano achieved strong automatic scores but showed isolated clinical inconsistencies. GLM-5 showed the most robust overall profile and performed close to human-authored notes in expert review.

**Conclusions:** LLMs can generate clinically plausible Spanish prostate cancer progress notes from published case reports under controlled conditions. These findings support the potential use of EHR-like synthetic corpora for clinical NLP, although reliability remains model- and prompt-dependent. Expert validation and safety-oriented evaluation are therefore necessary before downstream use or clinical deployment.

## 1 Introduction

### 1.1 Clinical Progress Summaries and Case Reports

Clinical progress notes, commonly referred to as evolution notes within Electronic Health Records (EHRs), are structured narrative documents that synthesize the longitudinal clinical status of a patient over time. This summaries plays a central role in healthcare, as they consolidate relevant clinical events, diagnostic findings and therapeutic decisions into an interpretable format. Their importance is particularly pronounced in the context of continuity of care [10], especially for patients with chronic conditions or complex oncological processes such as prostate cancer, where EHRs are critical for accurate clinical decision-making. In these scenarios, urologists and oncologists must interpret evolving data across multiple time points, making concise and reliable notes essential [26]. In parallel, another relevant source of clinical narratives is the clinical case reports published in the biomedical literature [14]. These documents describe individual patient cases, typically selected for their academic or educational value, rarity or clinical relevance. Unlike EHR-based notes, which are generated during routine care, case reports are retrospective, curated and often optimized for a didactic purpose. They tend to present a coherent and complete narrative, with explicit description of diagnostic reasoning, treatment decisions and outcomes. There are key differences between these types of clinical text. EHR notes are incremental, fragmented and written under times constraint, often containing abbreviations, omissions of irrelevant data and variability in structure [22]. In contrast, case reports are more structured, linguistically polished, and temporally coherent as they are written post hoc with full knowledge of the clinical trajectory. Overall, EHRs reflect real-world clinical practice and case reports provide cleaner and more interpretable narratives, albeit with potential biases due to selective reporting. However, modern clinical environments face a growing problem of documentation overload. The increasing volume of clinical data—ranging from laboratory results and imaging studies to treatment records—has made it progressively more difficult to extract and synthesize relevant information efficiently. This creates a tension between the need for comprehensive documentation and the practical constraints of clinical workflows. Two key dimensions highlight the importance of clinical notes. Firstly, the continuity of care and evolution of the clinical record, as clinical notes enable the integration of heterogeneous and temporally distributed information, including diagnostic procedures, therapeutic interventions and clinical events. By consolidating this information, they facilitate effective communication and handover between healthcare professionals. This continuity is directly linked to improved quality of care and enhanced patient safety, as it reduces the risk of information loss or misinterpretation [47]. Moreover, it reduces the administrative burden [41, 40], as a significant proportion of clinicians’ time is devoted to documentation tasks, often at the expense of direct patient care. This phenomenon, commonly described as *clinical documentation burden*, has been widely recognised as a major contributor to inefficiencies in healthcare systems. Excessive documentation requirements are also associated with increased levels of clinician burnout and reduced overall productivity [41]. Secondly, there is substantial inter-clinician variability in how medical evolution notes are written, including differences in style, level of detail and structural organization. This lack of standardisation leads to heterogeneous records, which can hinder interpretability and downstream processing. Furthermore, there is a temporal cost on manual generation of clinical evolution notes as it is inherently time consuming and does not scale well in high demand clinical environments. As patients volume increases, maintaining high quality documentation becomes increasingly challenging, further exacerbating the burden on healthcare professionals [40].

### 1.2 Large Language Models in Healthcare

Large Language Models (LLMs) have recently emerged as a transformative technology in the biomedical and clinical domains. These models, typically based on transformer architectures and trained on large scale textual corpora, demonstrate strong capabilities in natural language understanding and generation, making them particularly suitable for processing unstructured clinical text [42]. In this context, several key applications have been explored. Automatic summarization is possible given that LLMs enable abstractive summarization of clinical documents, allowing the generation of concise and coherent summaries from complex, contextualised, and heterogeneous sources such as EHRs or clinical case reports [4]. This is especially relevant for synthesising longitudinal patient information into structured evolution notes [23]. Clinical question answering is another application where LLM-based systems can be used, since they may be able to respond clinically oriented queries by extracting and synthesizing relevant information from patient records or biomedical literature [57]. These systems have been investigated as potential tools for supporting clinical decision-making and information retrieval [35]. Recent research has increasingly focused on the efficacy of LLMs in streamlining clinical documentation, with particular emphasis on their ability to generate high-fidelity narrativesincluding progress notes, discharge summaries, and case descriptionsdirectly from heterogeneous sources such as clinician-patient transcriptions [6]. This application directly targets the reduction of documentation burden by automating parts of the clinical writing process while maintaining medically meaningful content [20]. Despite the fact that all of these are promising capabilities, the deployment of LLMs in clinical settings introduces significant risks such as hallucinations, as LLMs may generate information that is not grounded in the input data. In clinical contexts, this can lead to the introduction of incorrect or fabricated details, such as non-existent diagnoses, mistaken laboratory values or treatments, which poses a substantial risk to reliability. In addition, generated outputs may exhibit internal contradictions, temporal inconsistencies, or violations of clinical logic. These issues are particularly problematic in longitudinal narratives, where maintaining coherence across time and events is essential for interpretability and safe clinical use [3].

### 1.3 Current Problem and Contribution

The problem addressed in this work is the automatic generation of synthetic clinical evolution notes from prostate cancer case reports published in the biomedical literature. The objective is not merely to summarize these reports, but to transform them into clinical narratives that resemble the evolution notes commonly found in hospital EHRs. This requires preserving, with strict factual accuracy, the critical clinical information contained in the original case report, including relevant diagnostic findings, procedures, treatments, disease status, temporal events, PSA values, Gleason scores and other clinically meaningful details. At the same time, the generated text must adopt the structure, level of detail, terminology, conciseness and narrative style characteristic of real-world clinical documentation. In this study, we focus specifically on the generation of synthetic clinical evolution notes for prostate cancer. To guide the expected style and clinical format of the generated narratives, we use as reference real evolution notes from the Urology Departments of two Spanish hospitals: Hospital Universitario Costa del Sol (HUCS) and Hospital Universitario Virgen de la Victoria (HUVV). Two of the authors of this study are clinicians working in these departments, which provides direct domain expertise regarding the structure, language and documentation practices of real hospital-based prostate cancer follow-up notes. Therefore, the main contribution of this work is the evaluation of LLMs for generating clinically realistic, EHR-like prostate cancer narratives that remain faithful to the source case report while resembling real-world hospital documentation. Building on this objective, the generated evolution notes must meet three essential requirements. First, they must remain faithful to the source case report, preserving the relevant clinical information while avoiding unsupported additions, omissions of critical facts or hallucinated content. Second, they must present the patients clinical trajectory in a coherent and temporally consistent manner, so that diagnostic findings, therapeutic interventions, disease evolution and outcomes are arranged in a clinically meaningful sequence. Third, they must be clinically interpretable and useful, meaning that the resulting narratives should resemble documentation that could support review, communication and decision-making in routine hospital practice. These requirements make the task more demanding than conventional text summarization. The model must not only generate fluent clinical language, but also transform a curated biomedical case report into an EHR-like evolution note while maintaining factual consistency, preserving clinically relevant details and reproducing the structure and style of real-world prostate cancer documentation. The present work makes the following main contributions:

- First, we provide a systematic comparative evaluation of LLMs for the generation of synthetic clinical evolution notes from published prostate cancer case reports. The analysis focuses on the ability of the models to preserve the clinically relevant information contained in the source case while producing narratives that resemble real-world hospital documentation. The comparison includes models with substantially different characteristics, including proprietary and open-source models, cloud-based and locally deployed models, and both general-purpose and biomedical domain-adapted language models. This allows us to assess not only overall generation quality, but also how different model families behave in a clinically constrained text generation task.
- Second, we analyse the impact of different prompting strategies on the quality and reliability of the generated clinical narratives. In particular, we examine how prompt design influences factual consistency, temporal coherence, structural organization and the preservation of critical prostate cancer information, such as diagnostic findings, treatments, PSA values and Gleason scores. This provides empirical evidence on the role of prompt engineering in clinical text generation and helps identify configurations that improve robustness and reduce unsupported or clinically inconsistent outputs.
- Third, we propose a multidimensional evaluation framework for assessing synthetic clinical evolution notes. This framework combines surface-level and semantic similarity metrics with clinically oriented criteria, expert clinical assessment and LLM-based evaluation. By integrating automatic metrics with domain-specific judgement, the proposed evaluation strategy enables a more comprehensive assessment of generated narratives, covering not only linguistic similarity but also clinical fidelity, usefulness, factual consistency and resemblance to real-world EHR documentation.

## 2 Related work

### 2.1 Clinical Text Summarization

Clinical text summarization is a long-standing task in biomedical NLP, motivated by the need to reduce the cognitive and temporal burden associated with the interpretation of large volumes of patient information, as shown by Sinsky (2016), which mentions that physicians might use up to 49.2% of their work time on EHR and desk work [53], In clinical practice, relevant information is often distributed across multiple notes, laboratory results, imaging reports, procedures, treatments, and follow-up encounters. Summarization methods aim to condense this heterogeneous information into concise representations that support clinical review, communication, and decision-making [43]. Early work in this area (Mishra et al, 2014) was mainly based on extractive approaches, where the system selected and reorganized salient fragments from the source documents. These methods offered an important advantage in terms of factual reliability, since the generated summaries were directly grounded in the original text [39]. However, they were limited in their ability to synthesize dispersed information, represent temporal progression and produce clinically coherent narratives that reflect the evolution of the patient over time [1, 9]. This limitation is particularly relevant in complex clinical domains, where clinically relevant information is often distributed across multiple sentences, notes, or encounters, and must be reconstructed through the integration of diagnostic findings, therapeutic decisions, longitudinal events, and clinical outcomes [48]. More recent work has therefore moved toward abstractive and hybrid summarization approaches. Rhazzafe (2024) sought not only to select information but also to reformulate, organise and integrate it into a coherent clinical narrative [50]. These approaches are better suited to longitudinal EHR data and structured clinical records, where the objective is often to produce a compact representation of a patient trajectory rather than a shortened version of a single document [23]. Nevertheless, clinical summarization remains especially challenging because the generated text must preserve medical accuracy, temporal consistency and clinical relevance. In addition, clinical narratives are frequently noisy, incomplete, repetitive and highly variable in style and structure, which complicates the development of robust summarization systems [60]. These challenges are directly related to the task addressed in this work, where the goal is to transform detailed prostate cancer case reports into synthetic clinical evolution notes that are concise, clinically coherent and faithful to the source case.

The introduction of LLMs has substantially changed the landscape of text summarization. Unlike earlier systems, LLMs can generate fluent and contextually rich summaries with limited task-specific supervision, making them particularly attractive for domains where annotated datasets are scarce or difficult to share [56, 4]. Two main strategies have been explored for adapting these models to summarization tasks: prompting-based generation and domain-specific fine-tuning. Prompting uses task instructions, constraints and examples to guide a pre-trained model toward the desired output format, offering flexibility and reducing the need for large labelled datasets. In contrast, fine-tuning adapts the parameters of a model to domain-specific corpora or tasks, potentially improving performance but requiring additional data, computational resources and careful control of overfitting or privacy-related risks [16]. In clinical NLP, these two paradigms are especially relevant. Prompting is useful when the objective is to impose explicit requirements on the generated summary, such as preserving critical facts, avoiding unsupported information, following a given clinical structure or adopting a specific documentation style [52] Research on clinical text summarization has evolved from extractive approaches toward more advanced abstractive methods. Early work primarily relied on selecting salient segments from clinical documents, which ensured factual consistency but failed to capture the underlying clinical reasoning and temporal evolution of patient [1] [9]. More recent approaches focus on integrating information across heterogeneous sources, including longitudinal EHR data and structured clinical records. A key challenge in this domain lies in preserving medical accuracy while generating coherent narratives that reflect the progression of clinical events. Additionally, the variability and noise inherent to clinical text remains significant obstacles for robust summarisation systems [60].

### 2.2 Large Language Models Summarization and Evaluation in NLP

LLMs have enabled a shift toward fully generative approaches to clinical summarization. In this context, two main paradigms have been explored: prompting-based methods and fine-tuning strategies. Prompting leverages the generalization capabilities of pre-trained models through task-specific instructions, offering flexibility and reduced data requirements. Meanwhile, fine-tuning adapts models to domain-specific corpora, potentially improving performance at the cost of increased resource demands [16]. Recent work has also explored reasoning-oriented prompting techniques, including chain-of-thought strategies, to enhance coherence and structure in generated outputs [58]. However, their application in clinical settings remains non-trivial, as increased reasoning steps may introduce inconsistencies or unsupported inferences, particularly in task requiring strict factual grounding.

### 2.3 Generation of Synthetic Clinical Text

The development of clinical NLP systems is traditionally hindered by the “privacy dilemma”, where access to real patient data is strictly restricted by regulations such as HIPAA and GDPR [12, 19, 51]. While de-identification is a standard prerequisite for data sharing, research has shown that even de-identified notes remain vulnerable to Membership Inference Attacks (MIA) [12, 51]. Consequently, synthetic clinical text generation has emerged as a robust alternative to mitigate privacy risks while addressing the critical shortage of shareable clinical corpora [18, 34, 51]. Early efforts in clinical text synthesis focused on structured data or short narratives using statistical and early neural architectures. Lee (2018) [27] utilized encoder-decoder models to generate chief complaints from discrete variables like age and diagnosis. Concurrently, Guan et al. (2018) proposed mtGAN [17], using Generative Adversarial Networks to synthesize electronic medical record (EMR) text [17, 28]. Progress continued with Melamud and Shivade (2019), who combined Long Short-Term Memory (LSTM) networks with Differential Privacy to generate clinical notes from the MIMIC-III database [36, 18]. Ive et al. (2020) expanded this to the mental health domain, employing Transformers to generate artificial mental health records guided by sense-bearing key phrases to maintain semantic coherence [19]. A significant shift occurred with the introduction of large-scale pre-trained models. Li et al. (2021) conducted the first systematic comparison of architectures—including CharRNN, SegGAN, GPT-2, and CTRL—for generating History and Present Illness (HPI) sections, demonstrating that GPT-2 was superior for downstream Named Entity Recognition (NER) tasks [28]. More recently, research has moved toward using LLMs like GPT-3.5 and GPT-4 for high-fidelity synthesis. Kweon et al. (2023) introduced Asclepius, the first publicly shareable clinical LLM trained on synthetic notes derived from PubMed Central case reports (PMC-Patients), demonstrating that literature-based synthesis can effectively replace sensitive hospital data [24]. Current literature emphasizes conditioned generation and the use of clinical guidelines. Ellershaw et al. (2024) pioneered the use of Royal College of Physicians (RCP) guidelines to prompt LLMs for the automated generation of structured discharge summaries [11]. Other studies have explored task-specific augmentation: Falis et al. (2024) and Bilioni et al. (2025) utilized LLMs to synthesize notes for automated medical coding (ICD assignment) [13, 5]. In specialized fields like radiology, Liu et al. (2025) and Pandita et al. (2025) demonstrated that local, open-source LLMs (e.g., Llama, Mistral) can achieve performance comparable to proprietary models in identifying limb fractures and converting dictations into structured reporting templates [34, 44]. In the Spanish clinical domain, contributions are growing to bridge the gap in non-English resources. Platas et al. (2024) proposed a hybrid method using real-world structured data from liver cancer cases to generate and automatically annotate Computed Tomography (CT) scan reports in Spanish, showing significant improvements in Named Entity Recognition (NER) performance when adding synthetic data to small training sets [46]. Additionally, Vakili et al. (2025) explored data-constrained synthesis for de-identification tasks across Swedish and Spanish, showing that moderately-sized LLMs can be domain-adapted with relatively small amounts of in-domain data to produce high-utility synthetic NER corpora [55]. These advancements suggest that synthetic generation is a viable pathway for expanding clinical datasets in Spanish and other resource-constrained languages. The evaluation of clinical text generation systems remains an open challenge. Traditional automatic metrics, such as BLEU and ROUGE [45, 33], are widely used but primarily capture lexical overlap, limiting their ability to assess semantic fidelity and clinical correctness. To address this limitation, recent work has explored embedding-based similarity measures and task-specific evaluation frameworks that better capture semantic alignment [59, 49]. Still, human evaluation continues to be the reference standard, particularly for assessing clinical validity and usefulness, although it is constrained given its high cost and scalability. More recently, the use of LLMs as evaluators (LLM-as-a-judge) has emerged as a complementary approach [21], enabling structured and multi-dimensional assessment of generated outputs. Despite their potential, questions remain regarding their reliability and alignment with expert clinical judgment, especially in high-stakes medical applications.

## 3 Methodology

### 3.1 Source Dataset and Clinical Entity Annotation

The dataset used in this study consists of 109 clinical case reports of prostate cancer patients written in Spanish and obtained from the biomedical literature. These documents provide detailed and structured descriptions of individual patient trajectories, including clinical presentation, diagnostic procedures, treatments, and outcomes. Their curated and retrospective nature makes them particularly suitable as a source for controlled clinical narrative generation task. In addition, as it is public data, there are no risk of data leakage, and allows the usage of cloud models. The search strategy was conducted using the MeSH descriptor “Prostatic Neoplasms” combined with the *Publication Type* ‘Case Reports’, applying filters for language (Spanish) and species (Humans). To increase the search sensibility, additional terms such as “Prostate Cancer” and “Adenocarcinoma/Carcinoma de Prostata” were included. The data sourced comprised primary databases—PubMed, SciELO, and LILACS—alongside complementary sources including Dialnet, Redalyc, and institutional repositories from Spanish-speaking universities. Publication inclusion criteria includes:

- Text should be individualized reports of human prostate cancer cases.
- Peer-reviewed articles, clinical notes, scientific letters, and academic theses describing specific cases.
- Full-text availability in Spanish. And exclusion criteria:
- Narrative or systematics reviews without individual patient descriptions.
- Non-disagreeable case series and experimental or preclinical (animal) studies.
- Duplicate publications of the same case and articles with no Spanish full-text available.

To support downstream analysis, a NER pipeline was applied to identify relevant clinical entities in the source texts, including diseases, symptoms, medical procedures, temporal expressions or other key clinical variables. The pipeline used consist of six NER models developed by the *Barcelona Supercomputing Center (BSC)*, and relies on the *BSC-NLP4BIA/PlanTL-GOB-ES* model family. These models are derived from the bsc-bio-ehr-es, a RoBERTa-based Spanish biomedical-clinical encoder pretrained on large-scale biomedical text and EHR data [8]. The following downstream token-classification models were used:

- BSC-NLP4BIA/bsc-bio-ehr-es-livingner-species/humano: fine-tuned for species and organism mentions using LivingNER resources and fine-tuned for human mention detection within the LivingNER framework [37].
- BSC-NLP4BIA/bsc-bio-ehr-es-medprocner: fine-tuned for medical procedure recognition in Spanish clinical texts on MedProcNER corpus [30].
- BSC-NLP4BIA/bsc-bio-ehr-es-symptemist: fine-tuned for symptoms, signs, and clinical findings using SympTEMIST resources [31].
- BSC-NLP4BIA/bsc-bio-ehr-es-distemist: fine-tuned for disease mention detection using DisTEMIST resources [38].
- PlanTL-GOB-ES/bsc-bio-ehr-es-pharmaconer: fine-tuned for pharmacological substances, compounds, and related biomedical mentions using PharmaCoNER resources [15].

In addition, three models from MedSpaNER family were used. These models are specifically designed for the semantic annotation of Spanish medical texts and cover clinical phenomena that are not fully addressed by the previous entity-recognition models [7]:

- medspaner/roberta-es-clinical-trials-cases-medic-attr: RoBERTa-based model for medication attributes, including dosage, route, form, and related drug information.
- medspaner/xlm-roberta-large-spanish-trials-cases-temp-ent: XLM-RoBERTa-large-based model for temporal entities, including dates, ages, durations, frequencies, and time expressions.
- medspaner/roberta-es-clinical-trials-cases-neg-spec: RoBERTa-based model for negation and speculation detection, including negation cues, negated concepts, speculation cues, and speculated events [32].

To complement the neural NER pipeline, two ad-hoc rule-based extractors were developed to identify PSA values and Gleason scores. These clinical variables are particularly relevant in prostate cancer, but they were not consistently captured by the selected NER models, mainly because they often expressed as numeric patterns, abbreviated mentions, or domain-specific scoring formats.

The PSA extractor relies on regular expressions to detect numeric values followed by standard PSA units, accounting for both decimal commas and decimal points, as well as common unit variants such as *ng/mL* or *ng/ml*. Similarly, the Gleason extractor captures several frequent formulations, including primary and secondary patterns such as *3+4*, total scores expressions such as *Gleason 7*, and combined formulations such as *Gleason 7 (3+4)* or *Gleason 3+4=7*.

These rule-based components were incorporated to improve the recall of prostate-cancer-specific variables that are clinically essential for staging, progression assessment, and treatment interpretation. Accordingly, they were used as complementary modules to the neural NER models, rather than as replacements.

On average, each document contains 68.32 annotated clinical entities, reflecting the high informational density of the source material. Since the corpus is derived from academic case reports, the frequency of misspellings and informal abbreviations is expected to be substantially lower than in authentic EHR documentation. Table B1 provides a detailed distribution of the annotated entity types. Procedures, symptoms, and diseases are the most frequent categories, which is consistent with the clinical narrative structure of prostate cancer case reports, where diagnostic procedures, clinical manifestations, and pathological conditions constitute the core informational content.

**Table 1:** Distribution of annotated entity types in the corpus.

| Entity type | Entities (n) | Entities (%) | Docs (%) |
| --- | --- | --- | --- |
| Procedure | 1559 | 20.9 | 100.0 |
| Symptom | 1170 | 15.7 | 100.0 |
| Disease | 1169 | 15.7 | 100.0 |
| Negated | 514 | 6.9 | 92.7 |
| Proteins | 504 | 6.8 | 94.5 |
| Negation cue | 476 | 6.4 | 92.7 |
| Human | 339 | 4.6 | 100.0 |
| Drug/chemical | 315 | 4.2 | 69.7 |
| Duration | 312 | 4.2 | 90.8 |
| Date | 241 | 3.2 | 72.5 |
| PSA | 196 | 2.6 | 69.7 |
| Age | 111 | 1.5 | 99.1 |
| Speculated | 111 | 1.5 | 51.4 |
| Speculation cue | 110 | 1.5 | 55.0 |
| Gleason | 93 | 1.2 | 71.6 |
| Frequency | 65 | 0.9 | 29.4 |
| Dose | 64 | 0.9 | 20.2 |
| Species | 51 | 0.7 | 13.8 |
| Route | 24 | 0.3 | 15.6 |
| Time | 20 | 0.3 | 12.8 |
| Unclear | 2 | 0.0 | 1.8 |
| **Total** | **7447** | **100.0** | **100.0** |

### 3.2 Constrained Generation of Synthetic Clinical Progress Summaries

The task is formulated as a constrained clinical text generation problem, where the primary objective is to produce a structured EHR-compliant evolution note from a comprehensive clinical case narrative. The input comprises a full-length clinical case report characterised by heterogeneous and temporally distributed information, including patient history, diagnostic procedures, therapeutic interventions, and clinical outcomes. The expected output is a concise, structured summary that accurately captures the most relevant aspects of the patients clinical trajectory while preserving the style, terminology, conciseness, and organisational conventions typically found in authentic EHR prostate cancer progress notes.

Unlike standard summarisation tasks, which primarily focuses on content selection and compression, this specific problem requires the rigorous preservation of clinical semantics and temporal coherence. To successfully emulate real-world evolution notes, the generated summaries must accurately reflect the chronological progression of events, ensuring absolute consistency across longitudinal diagnoses, interventions, and patient outcomes. Figure 1 illustrates this transformation process through a representative example. The left column shows an excerpt from a literature-derived prostate cancer clinical case, while the right column presents the corresponding synthetic EHR-style progress note generated from it (using GLM-5 with GS1 prompting strategy, see sections 3.3 and 3.4 below). Coloured spans indicate the clinical mentions automatically identified in each text by the NER models (see Section 3.1), including disorders, procedures, signs and symptoms, patient attributes, biomarker values, and contextual cues such as negation or speculation. This side-by-side representation makes explicit the central requirement of the task: the generated note must not merely compress the source narrative, but preserve the clinically relevant entities and their contextual interpretation while reorganising the information into a more concise and structured format. Furthermore, the generation process is bound by strict clinical constraints. The generated outputs must avoid safety-critical errors, most notably the introduction or hallucination of unsupported medical information. The system must also rigorously guard against internal inconsistencies, such as contradictory statements or deviations from established clinical logic. Given the longitudinal nature of oncological care, particular attention must be paid to preserving temporal consistency, as sequential disordering or misinterpretation of clinical events would compromise both the interpretability and reliability of the summary. Finally, the task implicitly involves handling of domain-specific terminology and high-density clinical information. The model must accurately represent complex key medical entities and their relational dependencies, alongside the ability to abstract and reorganise relevant content into a standardised format without omitting critical clinical details. Ultimately, by unifying abstractive summarisation, information extraction, and clinical reasoning under exacting reliability constraints, this problem stands as a particularly challenging task within the domain of clinical NLP.

**Figure 1:**
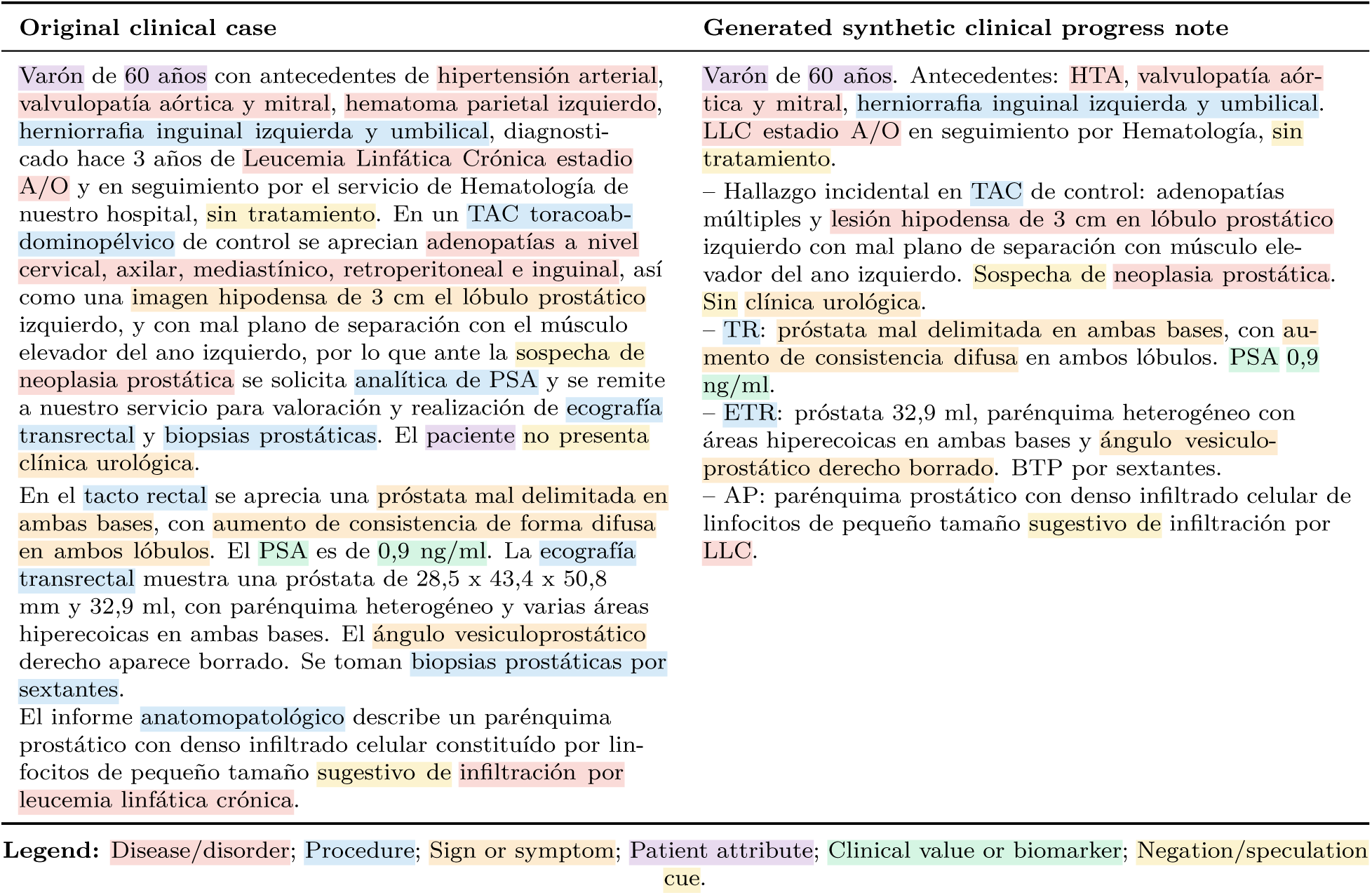
Illustrative transformation from a literature-derived clinical case into a synthetic elec-tronic health record progress note. Coloured spans denote automatically detected clinical mentions in both texts, highlighting the preservation of key entities and contextual modifiers such as negation and speculation during the generation process. (Texts are in Spanish. See complementary Figure C1 for the corresponding English translation.).

### 3.3 LLMs for Realistic Clinical Progress Note Generation

This study considers a wide variety of current state-of-the-art LLMs with different architectural paradigms, training strategies, and deployment settings, allowing for a heterogeneous comparison in the context of clinically-realistic synthetic text generation. The selected models include OpenAI GPT-5.4 Nano, Alibaba Qwen 3.5:35b, and GLM-5 as generative systems, alongside Claude Sonnet 4.6 as an evaluation model. A summary of the models used in this study is provided in Table 2..

- GPT-5.4 Nano (OpenAI) is a proprietary model, accessed via cloud-based APIs (Application Programming Interfaces). As a closed-source model, detailed information about its architecture and parameter count is not publicly available. It is designed to provide efficient inference while maintaining strong performance across general generation tasks, supporting a context window of up to 400,000 tokens and reasoning token capabilities.
- Qwen 3.5:35b was selected for local inference using its A3B variant, an optimised configuration that enhances inference performance. This LLM is designed for high-capacity language understanding and generation, as it supports multilingual inputs and exhibits strong performance in reasoning tasks. As an open-weights model, it can be deployed locally, mitigating the risk of data leakage and allowing greater control over inference settings such as temperature or seeding. In our specific case, the model was deployed on a dedicated GPU instance with 26GB of VRAM, achieving a mean latency of 5.36s per patient and a throughput of 73.76 tokens per second. This model exhibits strong performance in instruction-following settings, making it well-suited for prompt-based generation strategies such as zero-shot and few-shot learning. Its ability to adapt to structured prompts allows for controlled generation aligned with clinical constraints, including the preservation of key entities and temporal coherence [2].
- GLM-5 is an open-source, multilingual general-purpose model, based on a Mixture-of-Experts (MoE) architecture. This design enables dynamic routing of tokens across specialised expert networks, improving scalability and efficiency. It is designed to handle complex generation tasks across multiple languages, making it suitable for heterogeneous clinical narratives. Given its 744B parameters, inference was performed via cloud [61].
- In addition to the generative models, BioMistral 7B is included as a domain-adapted baseline [25]. This model is specifically trained on biomedical corpora and provides insight into the impact of domain specialisation compared to general-purpose LLMs. However, the model’s performance was suboptimal and fell significantly short of the quality standards established by the other models employed in this task. Due to its performance falling substantially below the established baseline for this task, the results for BioMistral 7B have been omitted from the subsequent analysis and comparative discussion.
- For evaluation purposes, a separate LLM was employed. In this study, a proprietary model was chosen—Claude Sonnet 4.6, by Anthropic—as an LLM-based evaluator to provide structured assessments across multiple clinically relevant dimensions. This separation between the generation and evaluation models reduces bias and enables a more robust analysis of output quality.

**Table 2:** LLMs for clinical progress summary generation and evaluation.

| Model | Role | Access | Notes | Parameters |
| --- | --- | --- | --- | --- |
| GPT-5.4 Nano | Generator | Cloud | Proprietary general model | Undisclosed |
| Qwen 3.5:35b | Generator | Local | Open-weights, controlled inference | 35b (3b windows) |
| GLM-5 | Generator | Cloud | Large Open Source MoE model | 744b |
| Claude Sonnet 4.6 | Evaluator | Cloud | Proprietary general model | Undisclosed |

*Note.* BioMistral 7B was also evaluated as a biomedical domain-adapted baseline, but its outputs did not meet the minimum quality requirements for this task and were therefore excluded from the comparative analysis.

### 3.4 Prompt Strategies

Two prompting strategies are used to guide the generation process (see Appendix D for the complete prompt templates). The first strategy, referred to as **Generation Strategy #1** and implemented through prompt **GS1**, uses the original clinical case report as plain text, without explicit annotation markup. The second strategy, referred to as **Generation Strategy #2** and implemented through prompt **GS2**, uses an entity-augmented version of the same input, in which relevant clinical mentions are explicitly marked with embedded XML medical-entity tags. Both strategies are evaluated under zero-shot and few-shot settings.

The structure of the prompts follows a consistent format across strategies. Each prompt includes: (i) a role specification, instructing the model to act as a clinical urologist; (ii) a set of explicit generation constraints, defining what information must be preserved or excluded; (iii) stylistic guidelines aligned with clinical narrative standards; and (iv) the input clinical case. In addition, in the few-shot setting, example pairs are inserted between the instructions and the target input. The key difference between GS1 and GS2 lies in the representation of the input. In GS1, the clinical case is provided as plain text. In GS2, the input is enriched with XML-like annotations that explicitly mark previously NER-detected clinical entities and provide additional structural signals to the model, facilitating the identification of clinically relevant information during generation. Overall, this setup enables the analysis of how input representation and prompt design influence the quality, consistency and clinical reliability of generated summaries.

In the zero-shot setting, models receive only task instructions and the input clinical case. These instructions are designed to enforce strict clinical constraints, including preservation of relevant information, avoidance of hallucinations, and adherence to the narrative style of real-world EHRs. The prompts explicitly constrain the generation process by specifying requirements related to temporal coherence, clinical relevance, and stylistic characteristics such as brevity, high information density, and use of standard medical abbreviations.

In the few-shot setting, the prompts are extended with two manually curated examples consisting of pairs of original clinical cases and their corresponding evolution summaries. These examples were produced by prostate cancer medical experts in a hospital environment (HUVV), ensuring that the target outputs reflect realistic clinical writing practices. The inclusion of expert-generated examples allows the models to better align with domain-specific conventions, particularly in terms of structure, terminology, and implicit prioritization of clinically relevant information. These examples are selected to balance contextual guidance and input length constraints, and are representative of typical prostate cancer clinical trajectories, including diagnosis, treatment, progression, and complications, thus ensuring that the LLMs are exposed to realistic patterns of longitudinal clinical evolution.

### 3.5 Evaluation Framework

Our evaluation framework is designed to rigorously assess the quality of the generated synthetic clinical progress notes from the dual perspectives of safety-criticality and clinical utility. This approach integrates a task-specific clinical safety filter with a multidimensional quality assessment based on the *PDQI-9* framework [54] and 5-point Likert-scale scoring [29]. Before any quantitative analysis, all generated summaries are subjected to a binary clinical safety screening. This primary gatekeeping stage is designed to intercept critical errors that could compromise patient safety or invalidate the clinical interpretation of the document. The screening specifically targets two categories of failure:

- Safety-critical errors: Defined as the presence of factually incorrect or potentially harmful clinical information, such as inappropriate interventions, incorrect dosages, or the misapplication of clinical protocols.
- Irreconcilable clinical contradictions: Referring to logically inconsistent statements within the generated summary, such as statements where the reported disease status is fundamentally incompatible with the described clinical progression.

If a generated summary meets either of these failure criteria, it is immediately categorised as *unsafe*. This binary safety label acts as a hard exclusion criterion and takes precedence over any other subsequent evaluation dimensions, ensuring that clinical reliability remains the primary requirement of the evaluation process. For generated notes that pass the initial safety screening, a multidimensional quality assessment is conducted using a standardised framework based on the PDQI-9 criteria. Each dimension is evaluated on a 5-point Likert scale, where 1 indicates *unacceptable quality* and 5 represents *optimal performance*. The evaluation is organised into two complementary components:

1. Comparative evaluation (relative to the original clinical case):

- Temporal correctness: Consistency in the chronological ordering and representation of clinical events.
- Precision: The absence of hallucinated or erroneous information not supported by the source case.
- Exhaustiveness: The comprehensive inclusion of all clinically significant information from the source case.
2. Independent evaluation (intrinsic quality of the generated summary):

- Clinical usefulness: The relevance and practical nature of the information for clinical decision-making.
- Organisation: The logical structure and overall coherence of the document.
- Comprehensibility: The clarity of expression and the appropriate application of clinical terminology.
- Conciseness: The absence of redundancy and superfluous information.
- Synthesis: The ability to effectively integrate and condense complex clinical data.
- Internal consistency: The absence of internal contradictions or logical fallacies within the text.

In addition to multidimensional quality assessment, a binary authorship classification task is defined to evaluate the realism and clinical plausibility of the generated notes. In this task, evaluators are presented with individual clinical progress notes and asked to classify each one as either *human-authored* or *model-generated*. The objective is not to assess factual consistency with the source case, but rather to determine whether the generated note is stylistically, structurally, and clinically indistinguishable from a realistic EHR progress note. This task therefore provides a complementary measure of synthetic note realism.

The overall evaluation process follows a hybrid approach, combining automatic metrics, LLM-as-a-judge assessments, and expert human evaluation. Automatic evaluation is first applied using similarity-based and semantic metrics to obtain an initial quantitative characterisation of the generated outputs. In parallel, the LLM-as-a-judge framework provides structured, multidimensional scores aligned with the aforementioned clinical criteria described above, as well as the binary authorship classification. Finally, human evaluation is conducted by two medical experts following a standardised annotation protocol, ensuring consistency across cases and evaluators. For this purpose, they evaluated 20 cases in a blinded setup, with no prior knowledge regarding whether the texts were human-authored or model-generated.

### 3.6 Experimental Configuration

The experiments were conducted using the corpus of 109 prostate cancer clinical cases, which served as the source data for all evaluated configurations. Those 109 clinical cases result in 7447 total clinical entities, of which 3956 are unique. All models were tested under the same input conditions to ensure comparability across settings. Generation parameters were controlled for each model to ensure consistency and reproducibility. For GPT-5.4 Nano, generation was configured with a maximum output length of 3000 tokens, medium reasoning effort, and low verbosity. For Qwen 3.5:35b, generation was performed using a maximum of 3000 tokens with temperature set to 0.2, top-p sampling at 0.9, top-k at 40, and a repetition penalty of 1.05. A fixed random seed was used to ensure reproducibility, and inference was executed in non-streaming mode. GLM-5 was evaluated using the same prompt configurations and experimental structure, although detailed control over generation parameters depended on the cloud-based interface. All outputs were generated in Spanish and followed a standardised extraction process based on explicit delimiters, ensuring consistent post-processing across models.

### 3.7 Comparison Dimensions

The experimental analysis is structured along three main dimensions to enable a comprehensive evaluation of model behaviour under different conditions:

- First, performance is compared across models, allowing the assessment of differences in generation quality associated with architectural design, scale, and deployment setting.
- Second, the impact of prompting strategy is analysed by comparing zero-shot and few-shot configurations. This dimension evaluates how the inclusion of example-based guidance influences the quality, consistency, and clinical reliability of the generated summaries.
- Third, results are analysed across generation settings, specifically GS1 and GS2. This comparison captures the effect of input representation, contrasting plain-text inputs with entity-enriched inputs, and its influence on the models ability to identify and preserve clinically relevant information.

## 4 Results

### 4.1 Automatic Evaluation: Lexical Similarity

Tables 3 and 4 present the results obtained using surface-level and semantic automatic evaluation metrics, respectively, after being evaluated against the original clinical cases:

- BLEU: measures n-gram precision between the generated note and the reference text, with a brevity penalty to discourage excessively short outputs.
- METEOR: computes an alignment-based similarity score using unigram matches, with penalties for fragmented or poorly ordered matches.
- N-gram cosine: represents texts as n-gram frequency vectors and computes their cosine similarity to quantify lexical overlap.
- ROUGE-1 F1: computes the F1 score of unigram overlap between the generated note and the reference text.
- ROUGE-2 F1: computes the F1 score of bigram overlap, capturing local phrase-level similarity.
- ROUGE-L F1: computes the F1 score based on the longest common subsequence, capturing sequence-level overlap while allowing non-contiguous matches.
- SBERT cosine: computes cosine similarity between sentence-level embeddings produced by Sentence-BERT, capturing semantic similarity beyond exact lexical overlap.
- BERTScore-P: computes contextual embedding-based precision, measuring how much of the generated content is semantically supported by the reference.
- BERTScore-R: computes contextual embedding-based recall, measuring how much of the reference content is semantically covered by the generated note.
- BERTScore-F1: computes the harmonic mean of BERTScore precision and recall, providing a balanced contextual similarity score.

**Table 3:** Surface similarity metrics for GS1 and GS2 reported as mean *±* standard deviation.

| GS1 |  |  |  |  |  |  |  |
| --- | --- | --- | --- | --- | --- | --- | --- |
| Model | Prompt | BLEU | METEOR | N-gram cosine | ROUGE-1 F1 | ROUGE-2 F1 | ROUGE-L F1 |
| GLM-5 | Zero | 13.559 ± 7.022 | 0.420 ± 0.085 | 0.572 ± 0.083 | 0.519 ± 0.110 | 0.309 ± 0.088 | 0.448 ± 0.119 |
| GLM-5 | Few | 13.740 ± 7.151 | 0.415 ± 0.083 | 0.574 ± 0.085 | 0.521 ± 0.103 | 0.315 ± 0.091 | 0.454 ± 0.109 |
| GPT5.4 | Zero | 16.468 ± 6.731 | 0.461 ± 0.080 | 0.624 ± 0.084 | 0.577 ± 0.099 | 0.350 ± 0.079 | 0.513 ± 0.102 |
| GPT5.4 | Few | **17.984 ± 6.543** | **0.472 ± 0.071** | **0.630 ± 0.081** | **0.593 ± 0.087** | **0.367 ± 0.077** | **0.530 ± 0.089** |
| Qwen3.5:35b | Zero | 10.366 ± 5.288 | 0.390 ± 0.073 | 0.515 ± 0.074 | 0.510 ± 0.098 | 0.268 ± 0.072 | 0.445 ± 0.107 |
| Qwen3.5:35b | Few | 10.794 ± 5.682 | 0.395 ± 0.072 | 0.534 ± 0.073 | 0.517 ± 0.096 | 0.284 ± 0.075 | 0.448 ± 0.104 |

| GS2 |  |  |  |  |  |  |  |
| --- | --- | --- | --- | --- | --- | --- | --- |
| Model | Prompt | BLEU | METEOR | N-gram cosine | ROUGE-1 F1 | ROUGE-2 F1 | ROUGE-L F1 |
| GLM-5 | Zero | 17.884 ± 9.107 | 0.473 ± 0.095 | 0.640 ± 0.083 | 0.570 ± 0.109 | 0.363 ± 0.102 | 0.504 ± 0.121 |
| GLM-5 | Few | 15.589 ± 8.966 | 0.441 ± 0.095 | 0.606 ± 0.086 | 0.549 ± 0.112 | 0.336 ± 0.102 | 0.485 ± 0.118 |
| GPT5.4 | Zero | 24.147 ± 8.034 | **0.544 ± 0.076** | **0.713 ± 0.072** | **0.669 ± 0.082** | **0.445 ± 0.082** | **0.606 ± 0.091** |
| GPT5.4 | Few | 22.929 ± 8.564 | 0.525 ± 0.094 | 0.701 ± 0.076 | 0.647 ± 0.108 | 0.429 ± 0.096 | 0.586 ± 0.113 |
| Qwen3.5:35b | Zero | 14.950 ± 6.225 | 0.455 ± 0.076 | 0.592 ± 0.067 | 0.581 ± 0.087 | 0.335 ± 0.076 | 0.526 ± 0.103 |
| Qwen3.5:35b | Few | **30.554 ± 30.183** | 0.515 ± 0.249 | 0.685 ± 0.184 | 0.633 ± 0.221 | 0.443 ± 0.276 | 0.550 ± 0.262 |

**Table 4:** Semantic similarity metrics for GS1 and GS2 reported as mean *±* standard deviation.

| GS1 |  |  |  |  |  |
| --- | --- | --- | --- | --- | --- |
| Model | Prompt | SBERT cosine | BERTScore-P | BERTScore-R | BERTScore-F1 |
| GLM-5 | Few | 0.642 ± 0.103 | 0.813 ± 0.032 | 0.813 ± 0.032 | 0.812 ± 0.033 |
| GLM-5 | Zero | 0.657 ± 0.132 | 0.814 ± 0.035 | 0.814 ± 0.035 | 0.812 ± 0.035 |
| GPT5.4-nano | Few | 0.719 ± 0.093 | **0.831 ± 0.032** | **0.831 ± 0.032** | **0.830 ± 0.032** |
| GPT5.4-nano | Zero | **0.726 ± 0.107** | 0.828 ± 0.032 | 0.828 ± 0.032 | 0.827 ± 0.032 |
| Qwen3.5:35b | Few | 0.651 ± 0.101 | 0.803 ± 0.032 | 0.803 ± 0.032 | 0.802 ± 0.032 |
| Qwen3.5:35b | Zero | 0.664 ± 0.119 | 0.796 ± 0.032 | 0.796 ± 0.032 | 0.795 ± 0.032 |

| GS2 |  |  |  |  |  |
| --- | --- | --- | --- | --- | --- |
| Model | Prompt | SBERT cosine | BERTScore-P | BERTScore-R | BERTScore-F1 |
| GLM-5 | Few | 0.677 ± 0.104 | 0.822 ± 0.036 | 0.822 ± 0.036 | 0.821 ± 0.036 |
| GLM-5 | Zero | 0.727 ± 0.119 | 0.834 ± 0.035 | 0.834 ± 0.035 | 0.833 ± 0.035 |
| GPT5.4-nano | Few | **0.805 ± 0.092** | 0.853 ± 0.034 | 0.853 ± 0.034 | 0.852 ± 0.035 |
| GPT5.4-nano | Zero | 0.795 ± 0.097 | **0.856 ± 0.031** | **0.856 ± 0.031** | **0.856 ± 0.031** |
| Qwen3.5:35b | Few | 0.777 ± 0.181 | 0.843 ± 0.090 | 0.843 ± 0.090 | 0.842 ± 0.090 |
| Qwen3.5:35b | Zero | 0.729 ± 0.119 | 0.820 ± 0.030 | 0.820 ± 0.030 | 0.819 ± 0.030 |

From a semantic perspective (Table 4), all models achieve consistently high similarity scores, with *SBERT cosine* and *BERTScore* values indicating strong alignment with the source clinical cases. GPT5.4-nano models obtain the highest scores across all semantic metrics in both GS1 and GS2, with GLM-5 closely following. Qwen 3.5:35b shows slightly lower performance, particularly under the GS1 setting, suggesting reduced semantic alignment when operating on plain-text inputs. Across strategies, GS2 generally leads to improved semantic similarity compared to GS1. This trend is consistent across models and metrics, indicating that the inclusion of entity-level information helps preserve clinically relevant content during generation. The improvements are particularly noticeable in *SBERT cosine* similarity and *BERTScore F1*, reflecting better global semantic coherence. In contrast, surface-level metrics (Table 3) show lower values and greater variability across models and configurations. This behaviour is expected given the abstractive nature of the task, where summaries involve compression, reorganization, and stylistic transformation of the original clinical case. As a result, lower *BLEU*, *ROUGE*, or *METEOR* scores do not necessarily indicate worse performance, but rather a higher degree of abstraction. When low surface similarity is accompanied by high semantic similarity it suggests that the generated summaries successfully preserve the underlying clinical meaning while applying appropriate abstraction. In this context, semantic metrics provide a more reliable signal of quality. The comparison between GS1 and GS2 shows a consistent increase in surface-level scores under GS2, indicating greater lexical alignment with the source text when entity-enriched inputs are used. However, this increase should not be directly interpreted as an improvement in the quality of the generated notes, as higher lexical overlap may reflect reduced abstraction rather than better clinical note.

### 4.2 LLM Evaluation

Figures 2 and 3 present the LLM-as-a-judge evaluation results in terms of both count-based outcomes and numerical scores across models, prompting strategies, and generation settings.

**Figure 2:**
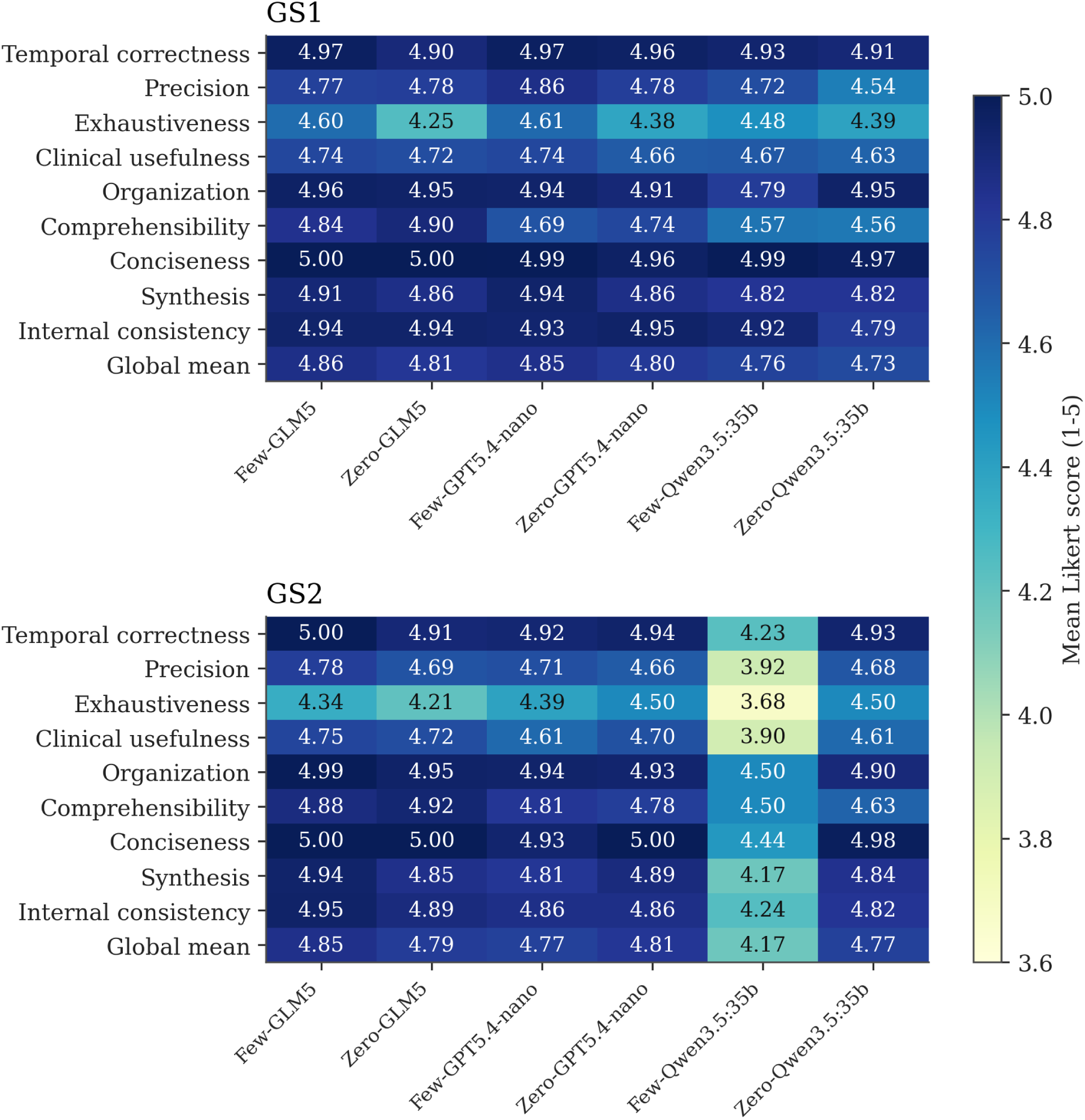
Claude Sonnet 4.6 Quantitative Evaluations for GS#1 and #2. Few and Zero refers to Few-shot experiments and Zero-shot experiments respectively.

**Figure 3:**
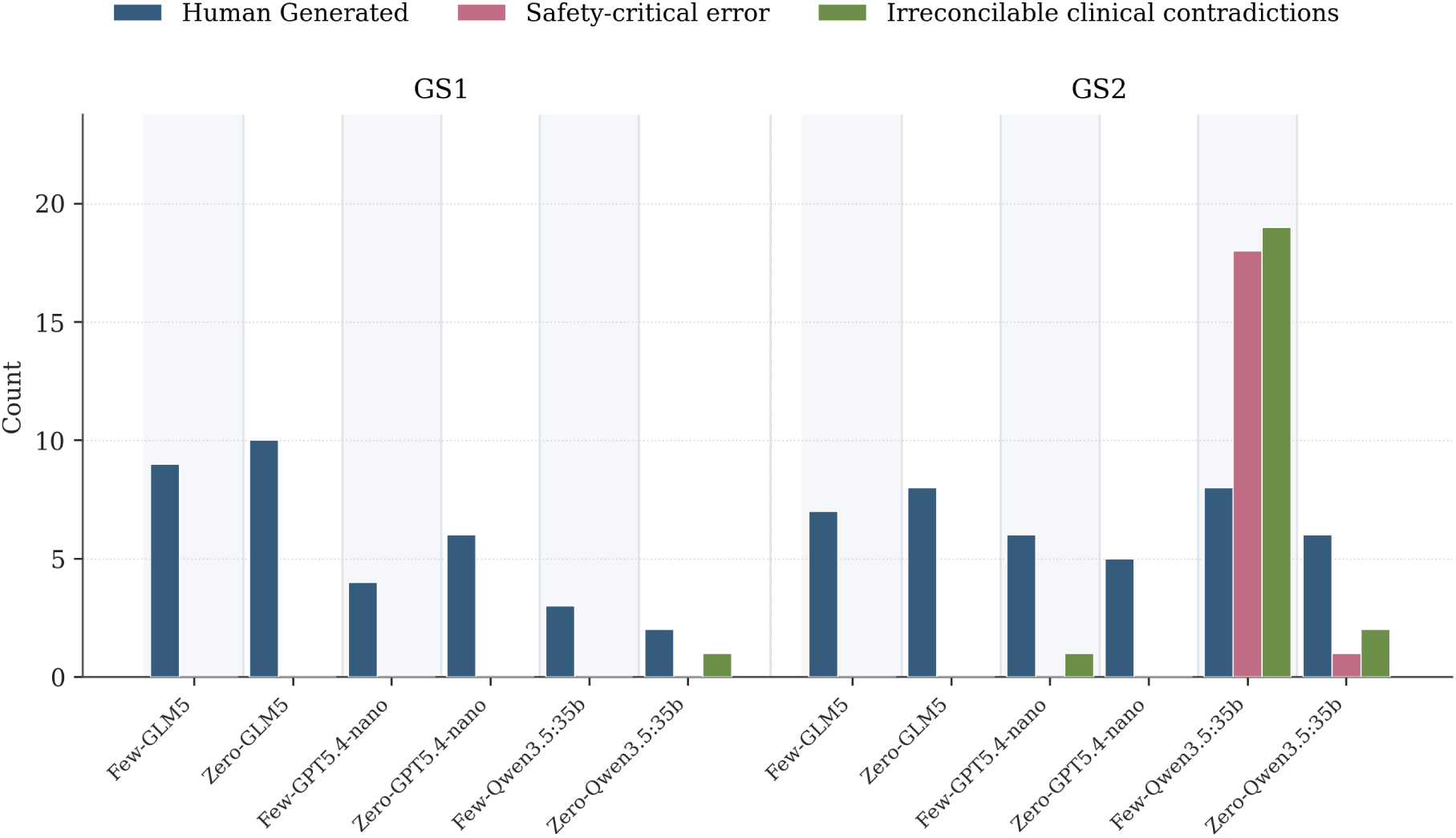
Claude Sonnet 4.6 Boolean Evaluations for GS#1 and #2. Few and Zero refers to Few-shot experiments and Zero-shot experiments respectively.

The numerical evaluation (Figure 2) provides a more granular view of performance across the nine PDQI-9 dimensions. In GS1, all models achieve consistently high scores across dimensions, with global mean values above 4.7 out of 5 on the Likert scale. GLM-5 slightly outperforms the other models, particularly in temporal correctness, organisation, and internal consistency. Differences between zero-shot and few-shot configurations are minimal in this setting, suggesting that prompting has limited impact when the input is provided as plain text.

From a safety perspective (Figure 3), GS1 shows consistently safe behaviour across all models, with no safety-critical errors and virtually no irreconcilable clinical contradictions. In contrast, GS2 introduces a clear degradation in safety for specific configurations. Notably, Qwen 3.5:35b under the few-shot setting exhibits a substantial increase in both safety-critical errors and clinical contradictions, representing the worst-performing configuration in terms of reliability. GPT5.4-nano show minor issues under GS2, while GLM-5 remains stable across both settings.

In GS2, performance becomes more variable. While GLM-5 and GPT5.4-nano models maintain relatively stable scores, Qwen 3.5:35b shows a significant drop across multiple dimensions, particularly in temporal correctness, precision, and clinical usefulness. This degradation is consistent with the increase in safety-related errors observed in the count-based analysis. The global mean score for Qwen 3.5:35b in the few-shot setting is notably lower than the rest of the configurations, indicating reduced overall quality when given a knowledge amplified input.

Across both settings, *conciseness* and *organisation* remain consistently high for all models, suggesting that stylistic constraints imposed by the prompts are effectively followed. On the contrary, *exhaustiveness* and *precision*exhibit greater variability, highlighting the difficulty of balancing completeness and factual accuracy in clinical summarization tasks.

Overall, the results indicate that while entity-enriched inputs (GS2) can provide additional structural information, they may also introduce instability depending on the model. In particular, local models such as Qwen 3.5:35b appear more sensitive to this input representation, especially in few-shot configurations. On the contrary, GLM-5 demonstrates the most robust performance across all conditions, maintaining both high-quality scores and strong safety performance. Given that professional clinical review is highly resource-intensive, evaluation was prioritised for the GS1, Few Shot experiment with the GLM-5 model. This selection stems from its higher mean score and its robustness, as evidenced by the lack of critical errors in any of the experimental trials

### 4.3 Clinical Assessment

The manual clinical assessment confirmed the strong performance of GLM-5 under the selected GS1 few-shot configuration (Figure 4 and Table 5). Across the 20 blinded review rows, GLM-5 achieved a global mean Likert score of 4.68, compared with 4.75 for the human-authored notes. Although reviewer R2 assigned lower scores than reviewer R1 for GLM-5, this pattern was not specific to model-generated text: the R2 global mean was identical for GLM-5 and human-authored notes, which indicates that the lower R2 scores mainly reflect reviewer-level calibration rather than a systematic degradation of the generated notes.

**Figure 4:**
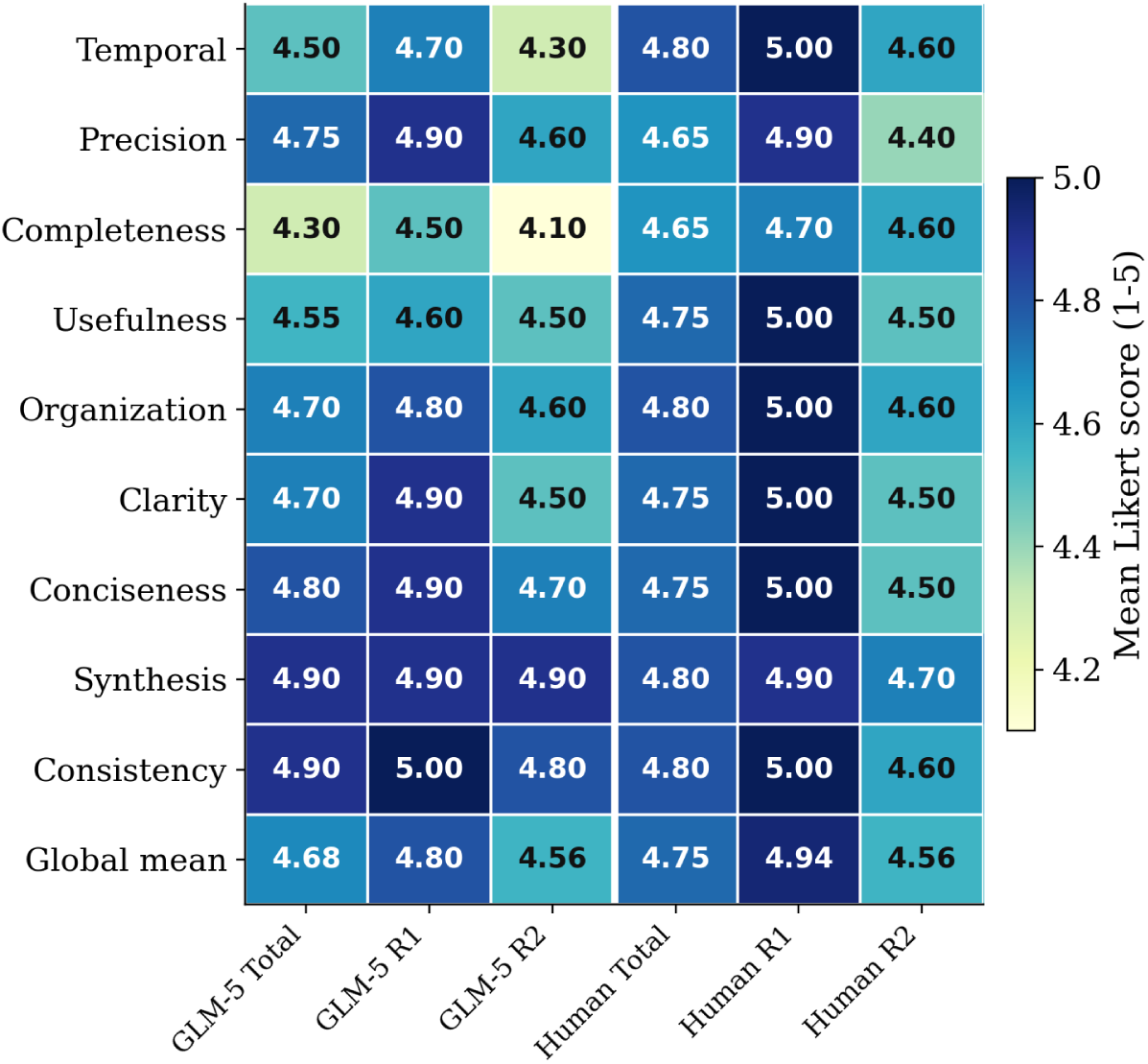
Manual clinical review of GLM-5 and human-authored notes across PDQI-9 dimensions. Scores are reported as mean Likert values on a 1–5 scale for both reviewers and globally.

**Table 5:** Manual review summary by generation source. Likert scores are reported as mean *±* SD on a 1–5 scale; categorical variables are reported as count and percentage of review rows. Excellent reports mean >4 and Adequate mean between 3 and 4. *HG means Human-generated.

| Source | Cases | R1 mean ± SD | R2 mean ± SD | Excellent | Adequate | Severe safety | Contradiction | HG yes | HG Unknown | HG no |
| --- | --- | --- | --- | --- | --- | --- | --- | --- | --- | --- |
| GLM-5 | 10 | 4.80 ± 0.41 | 4.56 ± 0.52 | 18/20 (90%) | 2/20 (10%) | 1 (5%) | 0 (0%) | 16 (80%) | 3 (15%) | 1 (5%) |
| Human | 10 | 4.94 ± 0.06 | 4.56 ± 0.44 | 19/20 (95%) | 1/20 (5%) | 0 (0%) | 0 (0%) | 19 (95%) | 1 (5%) | 0 (0%) |

The dimension-level analysis showed that GLM-5 remained close to the human-authored baseline across all PDQI-9 dimensions. The largest differences were observed in temporal correctness, completeness, usefulness, and clarity, whereas synthesis and consistency were nearly indistinguishable between GLM-5 and human-authored notes. Importantly, 90% of GLM-5 review rows were classified as excellent, compared with 95% for human-authored notes. In the blinded authorship task, 80% of GLM-5 notes were classified as human-generated and a further 15% as uncertain, while only 5% were explicitly classified as model-generated. These results suggest that most GLM-5 outputs were clinically and stylistically plausible enough to be judged as potentially human-authored.

### 4.4 Error Analysis

The error analysis focused on outputs explicitly flagged as containing either safety-critical errors or irreconcilable clinical contradictions, since these events represent the most relevant boundary for the potential clinical applicability of the system. Unlike minor issues related to style, conciseness, or exhaustiveness, these categories identify failures that may alter diagnostic interpretation, distort the clinical sequence, or introduce unsupported medical information. The stratified analysis by model and prompting strategy showed that these errors were not uniformly distributed. The highest concentration was observed for Qwen 3.5:35b under the GS2 few-shot configuration, which showed the most unfavourable risk profile and a marked overlap between safety-critical errors and irreconcilable clinical contradictions. This overlap suggests that the failures were not merely local inaccuracies, but rather structural deviations from the clinical representation of the source case. In comparison, the zero-shot configurations of Qwen 3.5:35b showed substantially fewer severe events, while GPT-5.4-nano under GS2 few-shot exhibited isolated cases of clinical contradiction without corresponding safety-critical errors. In the reviewed evaluation files, GLM-5 did not concentrate outputs marked as severe.

Clinical omissions were frequent, but they rarely constituted the main mechanism of potential harm. In most cases, omissions affected complementary information, including secondary comorbidities, complete laboratory values, technical details of procedures, histological nuances, or elements of the differential diagnosis. Although these losses reduce the exhaustiveness and traceability of the summary, they usually preserve the main diagnostic and therapeutic axis of the case. Their clinical relevance increases when the omitted information has an interpretative role, for example, in the assessment of disease progression, treatment toxicity, relevant comorbidity, or differential reasoning. Temporal inconsistencies had greater impact in outputs marked as clinical contradictions. The most relevant pattern was not a minor alteration in narrative order, but the generation of timelines that were incompatible with the original clinical evolution. In these cases, clinical milestones were displaced, inverted, or integrated into a sequence of care that could not be reconciled with the source case. This type of error is particularly critical in longitudinal clinical narratives, since temporality is not merely a formal property of the text, but a core component of clinical reasoning. It determines causality, disease progression, therapeutic response, and attribution of adverse events.

Hallucinations represented the most severe error category. In outputs marked as safety-critical, the generated information did not merely add undocumented accessory details. In several cases, it transformed structural elements of the case, including diagnosis, staging, biomarkers, therapeutic strategy, procedures performed, clinical evolution, or outcome. These hallucinations mainly followed two patterns. The first was factual reassignment, where a real clinical datum was attributed to the wrong entity, altering its medical meaning. The second was narrative substitution, where the generated summary no longer corresponded to the original patient and instead described an internally plausible but clinically false trajectory.

The distinction between irreconcilable clinical contradiction and safety-critical error is relevant for interpretation. Irreconcilable contradictions reflect internal incompatibilities or discordances with the source case that prevent the generated clinical note from being considered a faithful representation of the patient trajectory. Safety-critical errors, in contrast, identify outputs that could induce incorrect diagnostic or therapeutic decisions if used without expert review. In practice, both phenomena frequently overlapped: extensive hallucinations often produced incompatible timelines, incorrect diagnoses, and unsupported therapeutic plans simultaneously.

Overall, the error profile reveals a clear hierarchy of risk. Clinical omissions primarily affect completeness; temporal inconsistencies compromise longitudinal fidelity; and hallucinations, particularly when they reach the threshold of safety-critical error or irreconcilable clinical contradiction, directly compromise the clinical validity of the document. Therefore, it is confirmed that evaluation of clinical text generation systems should not rely exclusively on global quality scores, and should address direct comparison evaluation as LLM-as-a-judge or clinical expertise.

## 5 Discussion

The results support the viability of autonomous clinical progress note generation under controlled conditions, but also show that this feasibility is strongly model-dependent. The most relevant finding from the LLM-based evaluation is not only the difference in average quality scores, but the distribution of safety-critical failures across models, experiments and configurations.

As shown in Figures 2 and 3, GLM-5 achieves the most robust overall behaviour. Across the evaluated GS1 and GS2 configurations, no GLM-5 output was flagged as containing a safety-critical error or an irreconcilable clinical contradiction. This result is particularly important because these two categories define the practical boundary for autonomous use: while minor omissions, stylistic deviations, or reduced exhaustiveness may affect quality, safety-critical errors and unrecoverable contradictions directly compromise the clinical validity of the generated note.

This behaviour distinguishes GLM-5 from the other models. GPT-5.4-nano maintains generally high numerical scores, but isolated contradictions are observed in the GS2 setting. Qwen 3.5:35b shows a less stable profile, especially under GS2 few-shot, where the number of safety-critical errors and irreconcilable contradictions increases substantially. This suggests that the combination of entity-enriched input and few-shot examples may introduce model-specific instability, potentially leading to template-driven or insufficiently grounded generation.

From a clinical perspective, the manual review strengthens this interpretation. The absence of critical failures in GLM-5 is more relevant than small differences in mean quality scores and performed close to the human-authored baseline. Therefore, the results indicate that GLM-5 provides the strongest evidence of reliability within the evaluated experimental setting. The blinded authorship classification provides an additional indication of realism. Most GLM-5 outputs were classified as human-generated, and only a small minority were explicitly identified as model-generated. This suggests that the generated notes approximated the stylistic, structural, and clinical conventions expected in realistic progress notes. Therefore, the main distinction between GLM-5 and human-authored documentation was not global perceived quality, but residual clinical risk, as one GLM-5 review row was still flagged as containing a severe safety issue (related to an absence of previous diseases). This reinforces the need to interpret the apparent human-level performance as task-specific and conditional on expert validation, rather than as evidence of unrestricted autonomous clinical deployment.

The findings also reinforce the importance of evaluating clinical text generation beyond global similarity or average quality metrics. A generated summary can be concise, well structured, and semantically close to the source case while still containing a clinically unacceptable error. For this reason, safety-critical indicators must be treated as hard constraints when assessing the suitability of LLM-generated clinical documentation.

Overall, the results suggest that autonomous LLM-based generation of clinical progress notes is viable in constrained clinical domains when the selected model shows a stable safety profile. In this study, GLM-5 is the only evaluated model that combines high quality scores with the absence of both safety-critical errors and irreconcilable clinical contradictions, making it the most suitable candidate for further validation and deployment-oriented evaluation. However, given its large-scale architecture and associated computational requirements, practical deployment would require either a secure cloud-based solution compliant with clinical data protection standards, investment in local infrastructure capable of supporting reliable and scalable inference, or model-compression strategies such as quantization. The latter could reduce computational cost and facilitate local deployment, although its impact on clinical fidelity, temporal consistency, and safety-critical error rates should be specifically evaluated before adoption.

### 5.1 Limitations

This study has several limitations. First, the generated clinical progress notes are derived from clinical case reports rather than from real EHR data. Although case reports provide coherent and medically rich narratives, they differ from routine clinical documentation in style, structure, completeness, and reporting bias. Therefore, the results may not fully generalize to real-world EHR environments, where notes are often more fragmented, abbreviated, and heterogeneous.

Second, the dataset size is limited. The experiments were conducted on 109 prostate cancer clinical cases, which provides a controlled evaluation setting but restricts the diversity of clinical trajectories, writing styles, and edge cases represented in the analysis. Larger datasets covering broader urological and oncological scenarios and real clinical documentation would be necessary to assess the robustness of the proposed approach more comprehensively.

Third, the evaluation process depends partly on subjective assessment. Although the use of structured criteria, safety filters, and standardised scoring reduces variability, dimensions such as clinical usefulness, synthesis, organisation, and interpretability remain inherently judgement-dependent. In addition, LLM-as-a-judge evaluation may introduce model-specific biases and should not be considered a full substitute for expert clinical review.

Fourth, generation was restricted to a 1:1 ratio, focusing exclusively on the comparison between models and strategies. This decision was driven by budgetary constraints, as the utilization of cloud-based models requires pay-per-use API access. Furthermore, all of these models are recent releases, entailing a high cost-per-token that necessitated a more streamlined experimental scope. Finally, the study focuses on a constrained clinical domain and a specific generation task. The conclusions, particularly regarding the strong safety profile observed for GLM-5, should therefore be interpreted within this experimental setting. Further validation would be required before extending the approach to other diseases, document types, hospitals, or deployment environments.

## 6 Conclusion

This work demonstrates the potential of LLMs for the automatic generation of clinical progress notes from prostate cancer case narratives. The task requires more than textual compression, as generated summaries must preserve clinical meaning, temporal coherence, and factual consistency while adopting a concise electronic-health-record style. In the manual blinded evaluation, GLM-5 reached a quality level close to the human-authored baseline, with very similar global Likert scores and most generated notes being classified as human-generated or indistinguishable from human-generated text.

A central finding of this study is that published clinical case reports can serve as a valuable source for generating EHR-like synthetic clinical documentation. Since these reports are publicly available and do not involve the same confidentiality restrictions as real patient records, they provide a practical and legally less constrained starting point for clinical text generation. The results show that current LLMs are able to produce high-quality synthetic notes that preserve key diagnoses, procedures, temporal information, biomarker values, and clinically critical details, while approximating the structure, terminology, and conciseness of real-world prostate cancer progress notes.

The findings also highlight the importance of evaluating clinical generation systems beyond conventional similarity metrics, incorporating safety-oriented criteria and clinically meaningful quality dimensions. In this context, robust model selection emerges as a central requirement for any future use of synthetic clinical text in downstream NLP and AI pipelines. In particular, the generation of realistic and clinically faithful synthetic corpora may help reduce the dependence on large volumes of real EHR data, facilitating the development of models for information extraction, entity recognition, clinical summarisation, temporal reasoning, and other downstream tasks under data-scarce or privacy-constrained conditions.

Future work should focus on scaling up the generation process and empirically validating the utility of these synthetic corpora for training downstream clinical NLP models. A key next step will be to assess whether models trained or pre-trained on EHR-like synthetic notes can improve performance on real-world clinical data, especially when access to large annotated EHR corpora is limited. Additional work should also address clinical fine-tuning, validation on real-world EHR data, longitudinal evaluation across more complex patient trajectories, and deployment strategies involving secure cloud inference, local infrastructure, and quantized models, while preserving clinical reliability. Under the controlled conditions evaluated in this study, GLM-5 provides evidence that LLM-generated clinical progress notes can reach a human-comparable baseline for this specific prostate cancer summarisation task, although residual safety risks still require expert oversight before any clinical use.

## Data Availability

All data produced are available online at Zenodo: 10.5281/zenodo.20303982

https://zenodo.org/records/20303982

## Data and Code Availability

All 109 clinical cases and all generated outputs can be found in Complete Dataset. For replicability purposes, all developed code has been upload to the following GitHub repository: Synthetic GenEHR. Please be aware that code execution might need high computational resources or a paid subscription

## Acknowledgments

The authors acknowledge the support from the Ministerio de Ciencia e Innovación (MICINN) under project PID2024-155334OB-I00.

## A Search protocol and eligibility criteria

### A.1 Study design

An observational descriptive study was designed to construct a curated clinical database of Spanish-language prostate cancer case reports. The aim was to identify individual published human cases without restrictions on publication year, country of origin or publication format, provided that the report contained patient-level clinical information.

### A.2 Operational definition of case report

A case report was operationally defined as any publication describing at least one individual patient diagnosed with prostate cancer and reporting direct clinical information on presentation, diagnosis, treatment, disease course or outcome. Case series were considered eligible only when each patient could be assessed separately.

### A.3 Inclusion criteria

Publications were included when they met all of the following criteria:

- Description of one or more patients diagnosed with prostate cancer.
- Individualised patient-level information.
- Full text available in Spanish.
- Direct clinical information on presentation, diagnosis, treatment, disease progression and/or outcome.
- Human subjects.
- Publication in any of the following formats:

**–** Peer-reviewed scientific articles.
**–** Local or regional clinical journals.
**–** Scientific letters.
**–** Clinical notes.
**–** Image-based case sections.
**–** Institutional university repositories.
**–** Academic theses, provided that specific clinical cases were described.

### A.4 Exclusion criteria

Publications were excluded when they met any of the following criteria:

- Narrative or systematic reviews without individual patient descriptions.
- Case series in which individual patients could not be separated.
- Experimental, preclinical or animal-model studies.
- Duplicate reports of the same clinical case.
- Articles for which the full text was not available in Spanish.

### A.5 Complementary manual search procedures

In addition to conventional database searches, complementary retrieval strategies were used to maximise sensitivity. These included manual searches in Spanish-language urology journals, screening of institutional repositories and structured searches in Google Scholar using exact Spanish expressions commonly found in case reports, such as “cáncer de próstata” “caso clínico”, “adenocarcinoma de próstata” “caso” and “se presenta el caso” “próstata”.

### A.6 AI-assisted search

A complementary AI-assisted search was performed using Gemini Deep Research in reasoning mode. The model was prompted to identify published Spanish-language prostate cancer case reports, provide literal case descriptions and generate bibliographic references in BibTeX format. Retrieved references were imported into Paperpile to support automatic PDF identification. All candidate records obtained through this route were manually reviewed before inclusion.

### A.7 Software-assisted search

A software-assisted search was also implemented using predefined prostate cancer terms, case-report terms and Spanish-language filters. Candidate records were prioritised using textual signals suggestive of individual case descriptions, including expressions such as “presentamos el caso”, “se presenta el caso”, “caso clínico”, “reporte de caso”, “serie de casos”, “varón de \d{2}” and “paciente de \d{2}”. Candidate references were exported in BibTeX format and processed through the same PDF retrieval and manual review workflow.

Then, a semi-automated abstract-screening step was used to prioritise candidate records. Titles and abstracts were evaluated with local LLMs, primarily qwen2.5:14b-instruct and, when unavailable, llama3.1:8b-instruct. The models classified records as eligible, non-eligible or uncertain according to the predefined criteria. All records classified as eligible or uncertain underwent manual full-text verification before inclusion in the final corpus.

Records were screened in two stages. First, titles and abstracts, or title and snippet in the case of Google Scholar records, were reviewed. Second, potentially eligible publications were assessed in full text to confirm compliance with the inclusion and exclusion criteria. Records with uncertain eligibility were retained for full-text review.

## B Named Entity Recognition

### B.1 Model-level details of the NER resources

The named entity recognition pipeline combined Spanish biomedical-clinical transformer models from the *BSC-NLP4BIA / PlanTL-GOB-ES* ecosystem with complementary MedSpaNER components. The objective was to extract structured clinical information from Spanish reports, covering diseases, symptoms, procedures, medications, living beings, temporal expressions and contextual phenomena such as negation or speculation.

*LivingNER* was used to detect mentions of living beings and biologically relevant entities. This includes references to humans, patients, relatives, healthcare professionals, pathogens, microorganisms and other species. The model is based on bsc-bio-ehr-es and was trained on Spanish clinical case reports annotated with species and human mentions.

*MedProcNER* was used to identify clinical procedures. It detects diagnostic tests, therapeutic interventions, surgeries and other healthcare-related actions described in the reports. The model is based on bsc-bio-ehr-es and was trained on Spanish clinical cases annotated with procedure mentions, often normalised to SNOMED CT.

*SympTEMIST* was used to recognise symptoms, signs and clinical manifestations. Its purpose was to capture patient-reported symptoms, observed signs and relevant clinical findings. The model uses bsc-bio-ehr-es and was trained on Spanish clinical case reports annotated by clinical experts, with SNOMED CT normalization.

*DisTEMIST* was included to detect diseases and diagnostic expressions. It identifies mentions of disorders, diagnoses and pathological conditions in Spanish clinical text. The model is based on bsc-bio-ehr-es and was trained on manually annotated clinical cases with disease mentions mapped to SNOMED CT concepts.

*PharmaCoNER* was used to recognise pharmacological entities. It detects drugs, active substances, chemical compounds and medication-related biomedical mentions. The model belongs to the PlanTL-GOB-ES ecosystem, uses bsc-bio-ehr-es, and was trained on Spanish clinical cases annotated by biomedical and clinical experts.

The negation and speculation component was used to identify whether clinical mentions were asserted, negated or uncertain. This is relevant because a disease or symptom mention may refer to an absent or suspected condition rather than a confirmed finding. This MedSpaNER component used Spanish medical corpora annotated for negation and uncertainty phenomena.

The temporal entity recognition model was used to detect time-related expressions. It identifies dates, durations, frequencies, times and age-related temporal references. This component, based on bert-base-multilingual-cased, added temporal context to the extracted clinical entities.

The medication attribute recognition model was used to extract details associated with medication use. It captures attributes such as dose, route, frequency, duration, dosage form and administration mode. This MedSpaNER component complemented *PharmaCoNER* by extracting treatment-administration information rather than only drug names.

Together, these models provided a broad entity-level representation of the clinical reports, enabling downstream analysis and generation to account for clinical entities, their context and their temporal or pharmacological attributes.

**Table B1:** Distribution of annotated entity types in the corpus, where Entities refers to the total of entities detected, % of entities to the proportional part of that type of entity across all detected and % Docs the percentage of corpus documents containing those entity type mentions.

| **Entity type**<br>**(n)**<br>**(%)**<br>**(%)** | **Entities**<br>**Entities**<br>**Docs** |  |  |
| --- | --- | --- | --- |
| Procedure | 1559 | 20.9 | 100.0 |
| Symptom | 1170 | 15.7 | 100.0 |
| Disease | 1169 | 15.7 | 100.0 |
| Negated | 514 | 6.9 | 92.7 |
| Proteins | 504 | 6.8 | 94.5 |
| Negation cue | 476 | 6.4 | 92.7 |
| Human | 339 | 4.6 | 100.0 |
| Drug/chemical | 315 | 4.2 | 69.7 |
| Duration | 312 | 4.2 | 90.8 |
| Date | 241 | 3.2 | 72.5 |
| PSA | 196 | 2.6 | 69.7 |
| Age | 111 | 1.5 | 99.1 |
| Speculated | 111 | 1.5 | 51.4 |
| Speculation cue | 110 | 1.5 | 55.0 |
| Gleason | 93 | 1.2 | 71.6 |
| Frequency | 65 | 0.9 | 29.4 |
| Dose | 64 | 0.9 | 20.2 |
| Species | 51 | 0.7 | 13.8 |
| Route | 24 | 0.3 | 15.6 |
| Time | 20 | 0.3 | 12.8 |
| Unclear | 2 | 0.0 | 1.8 |
| **Total** | **7447** | **100.0** | **100.0** |

## C Synthetic Generated Notes Examples

Please note this examples correspond to the GS1 Few-Shot strategy, generated by GLM-5

**Figure C1:**
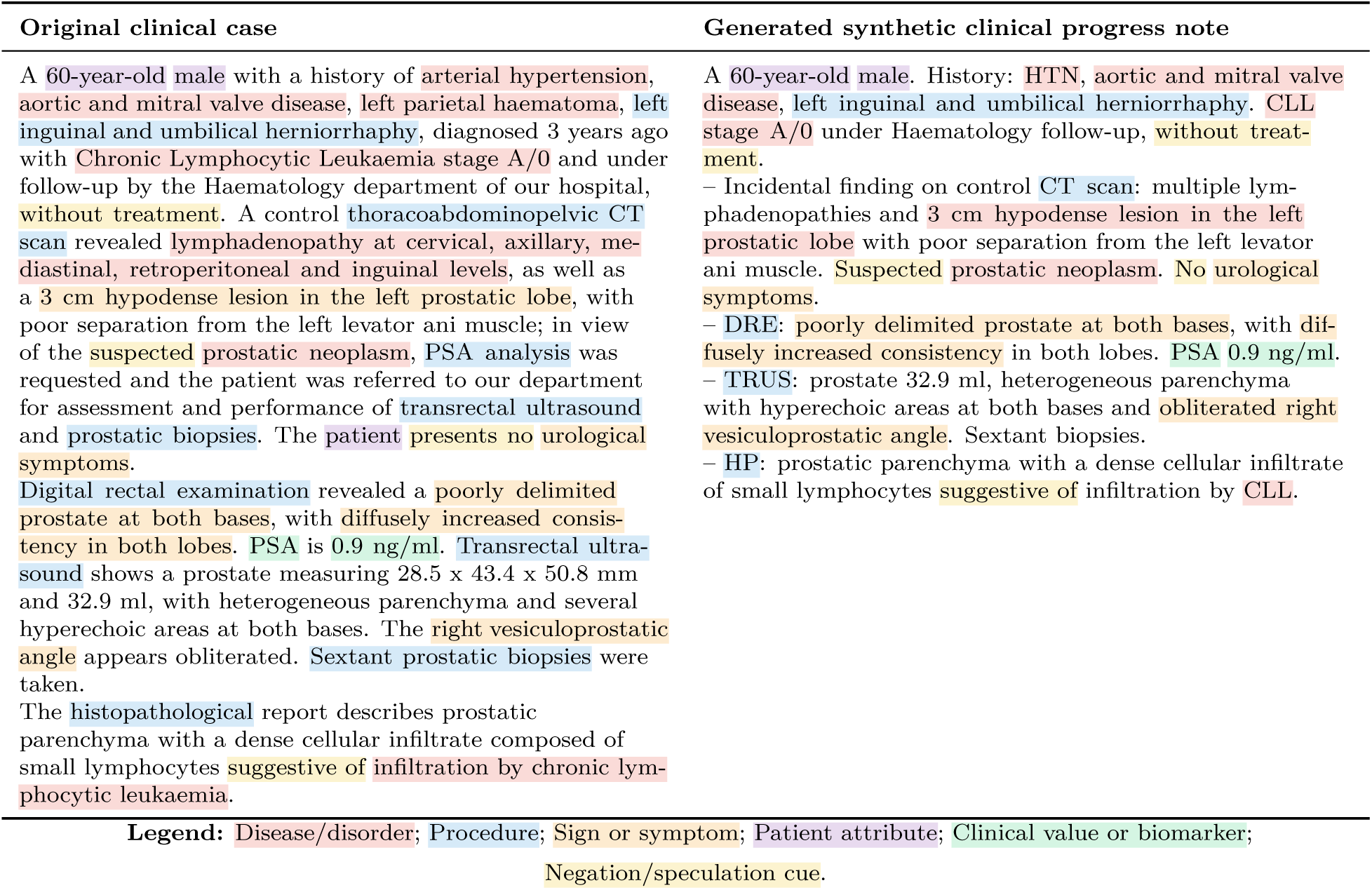
Illustrative transformation from a literature-derived clinical case into a synthetic electronic health record progress note. Coloured spans denote automatically detected clinical mentions in both texts, highlighting the preservation of key entities and contextual modifiers such as negation and speculation during the generation process.

## D Prompts

### D.1 GS1

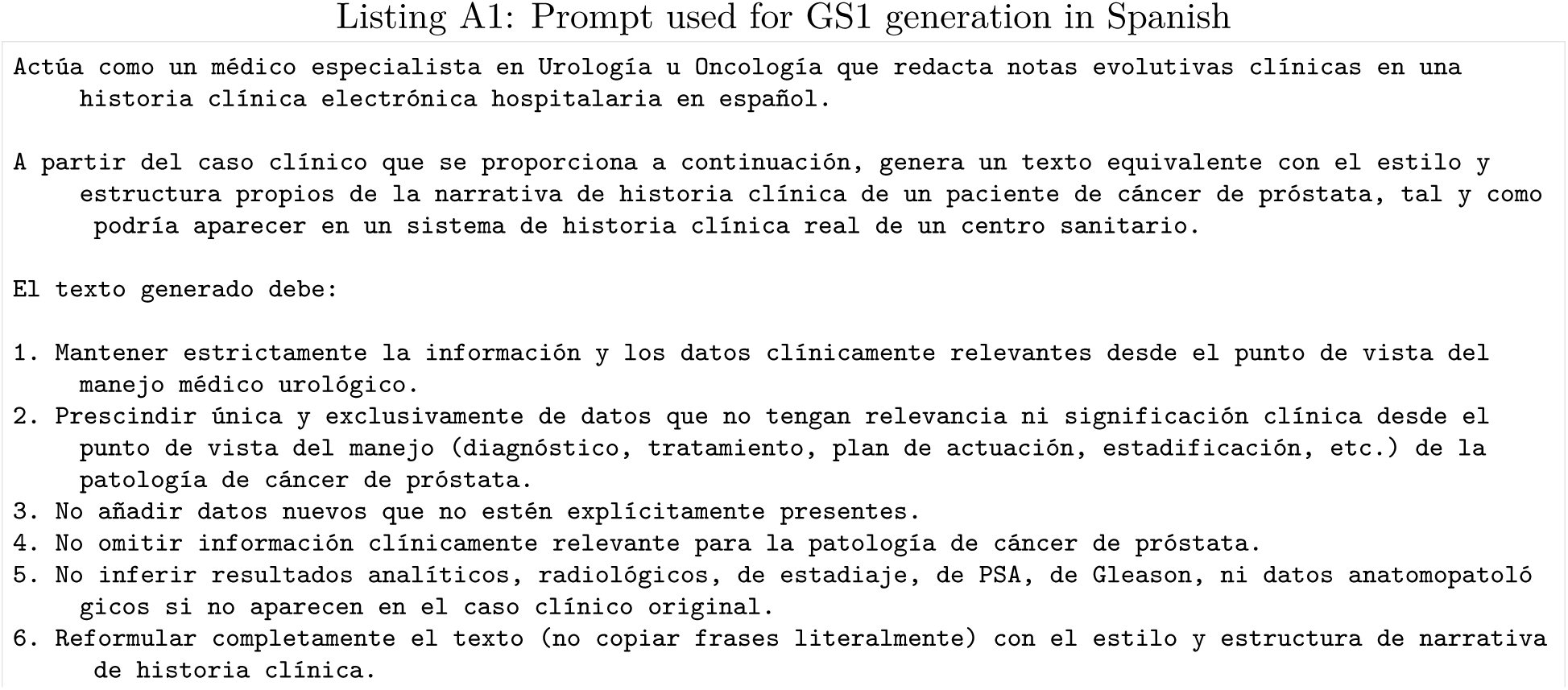
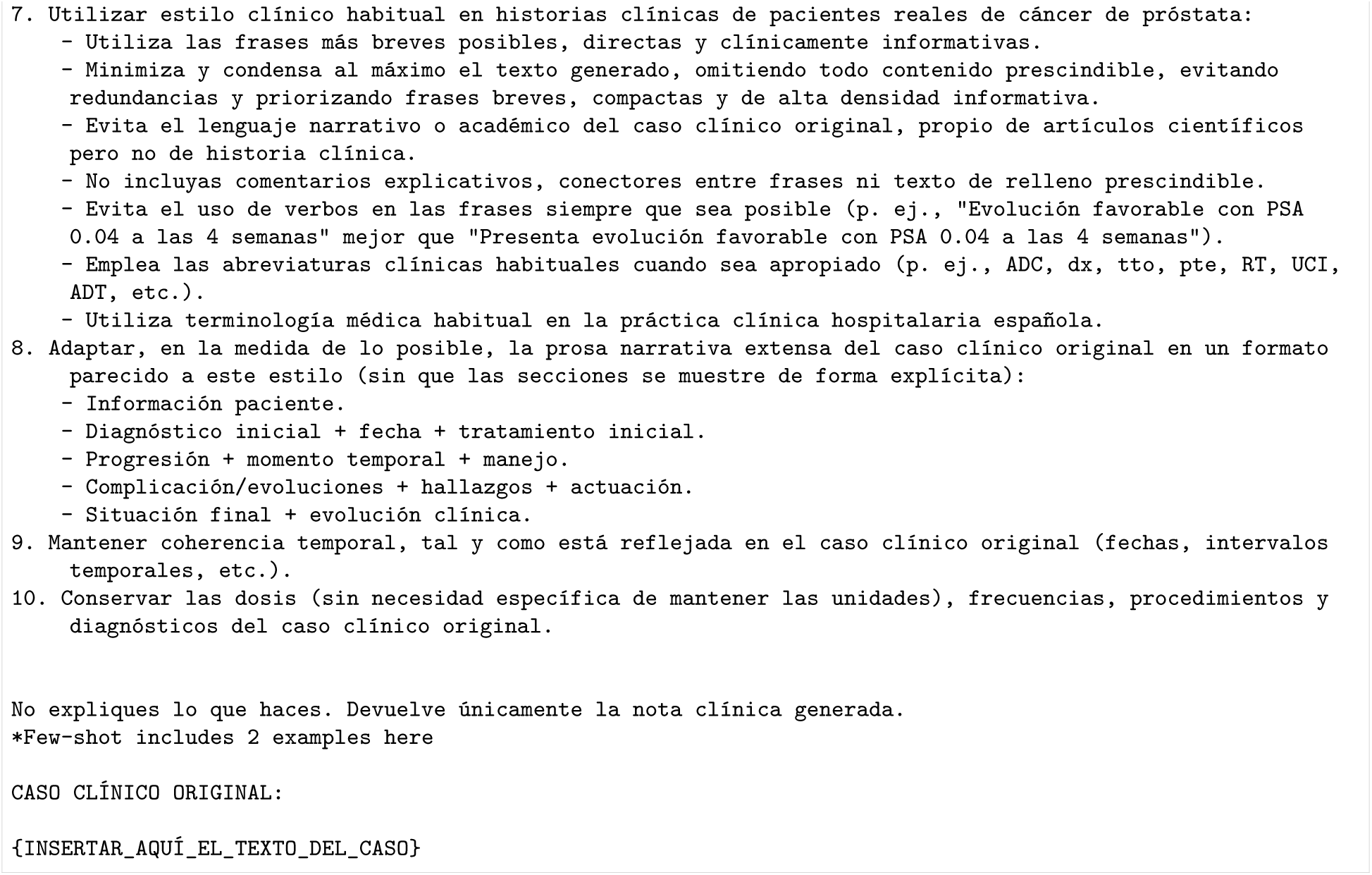

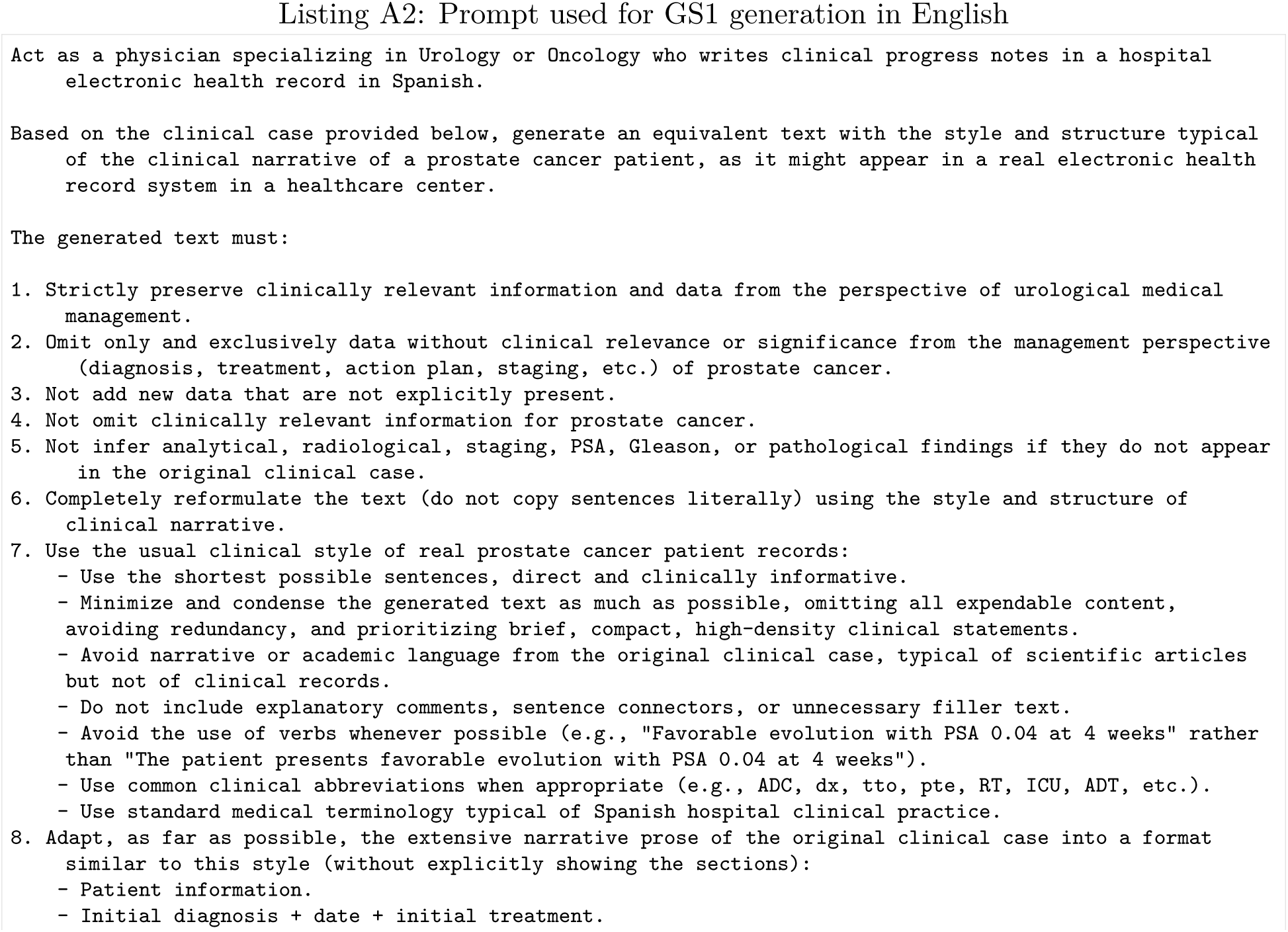
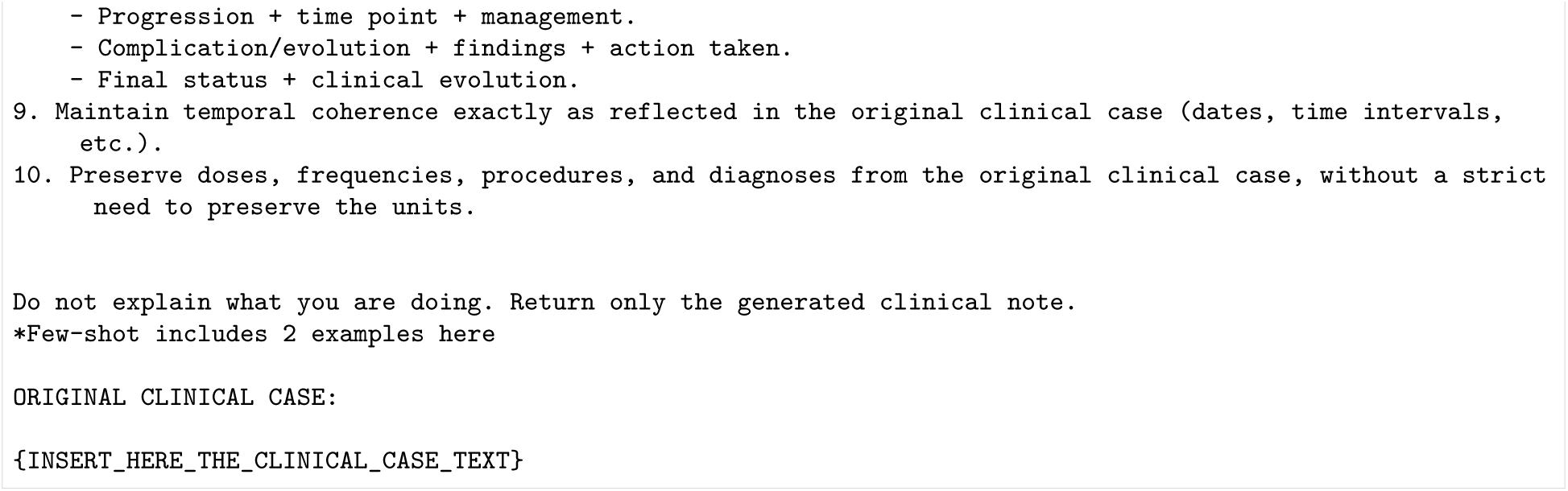

### D.2 GS2

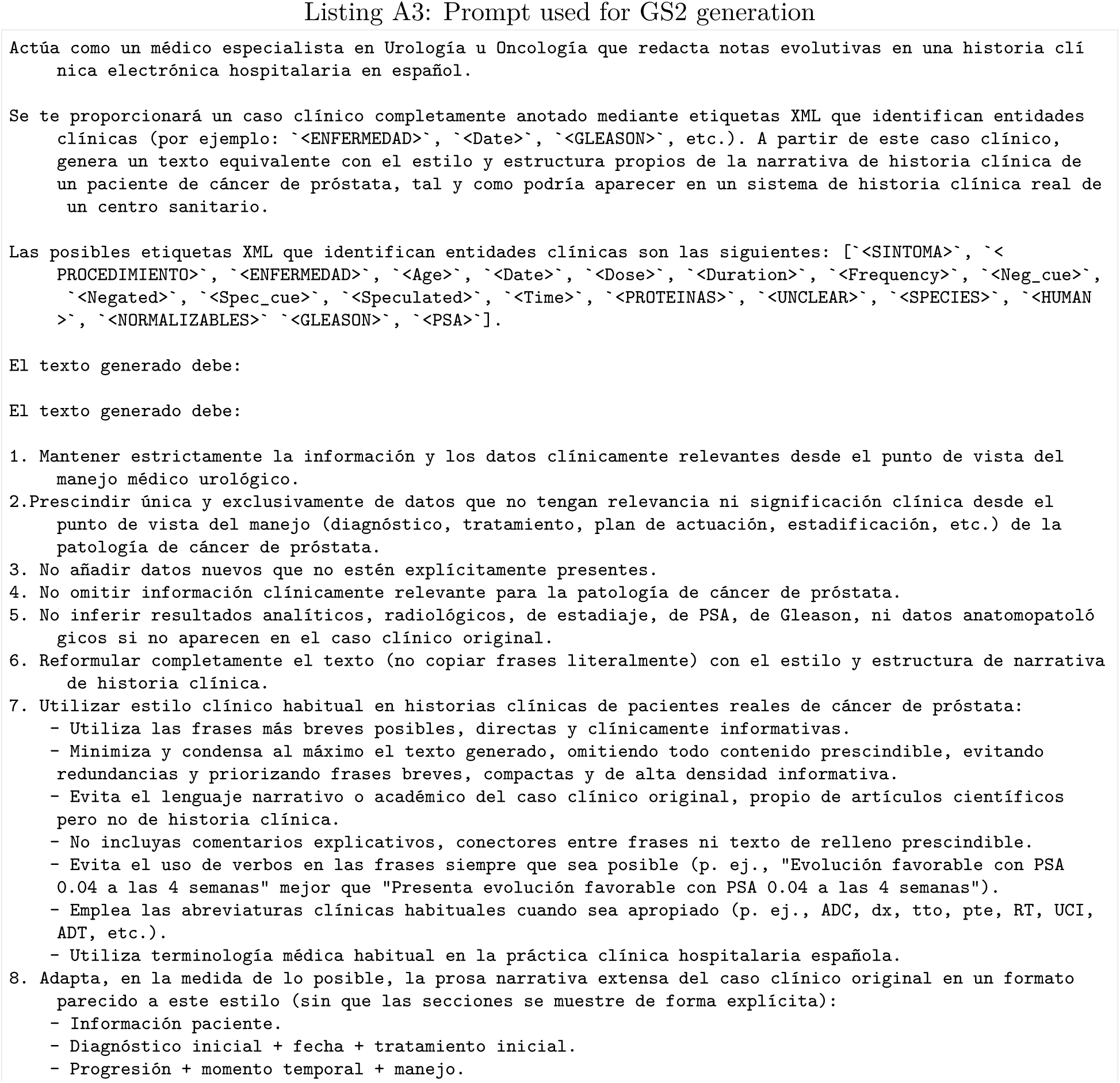
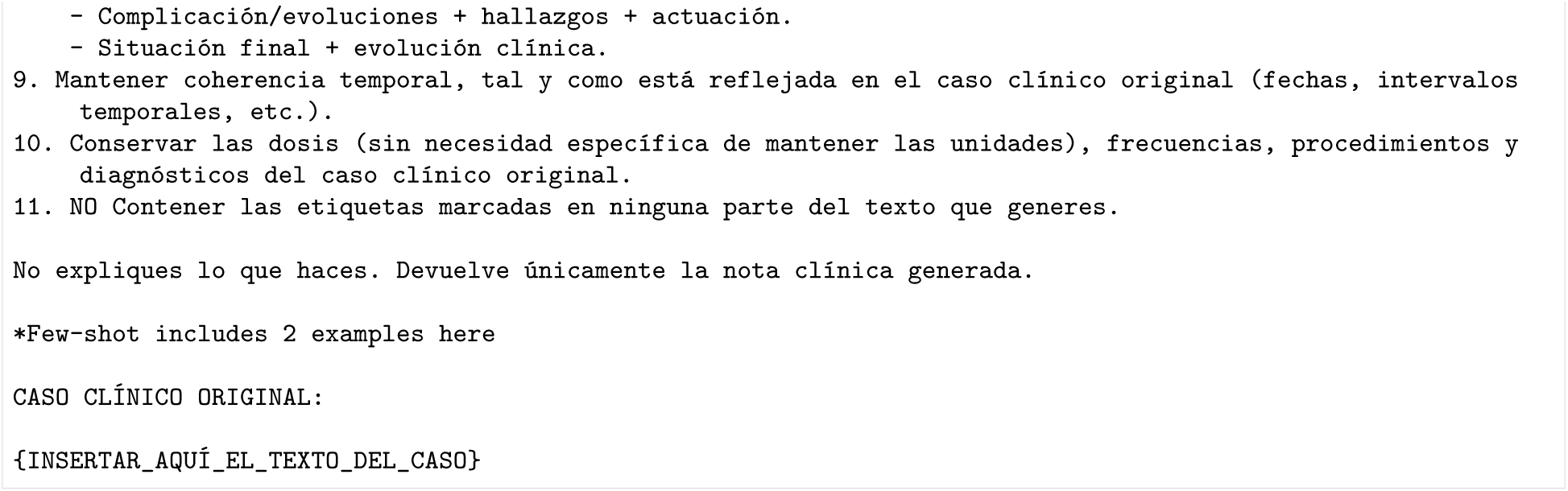

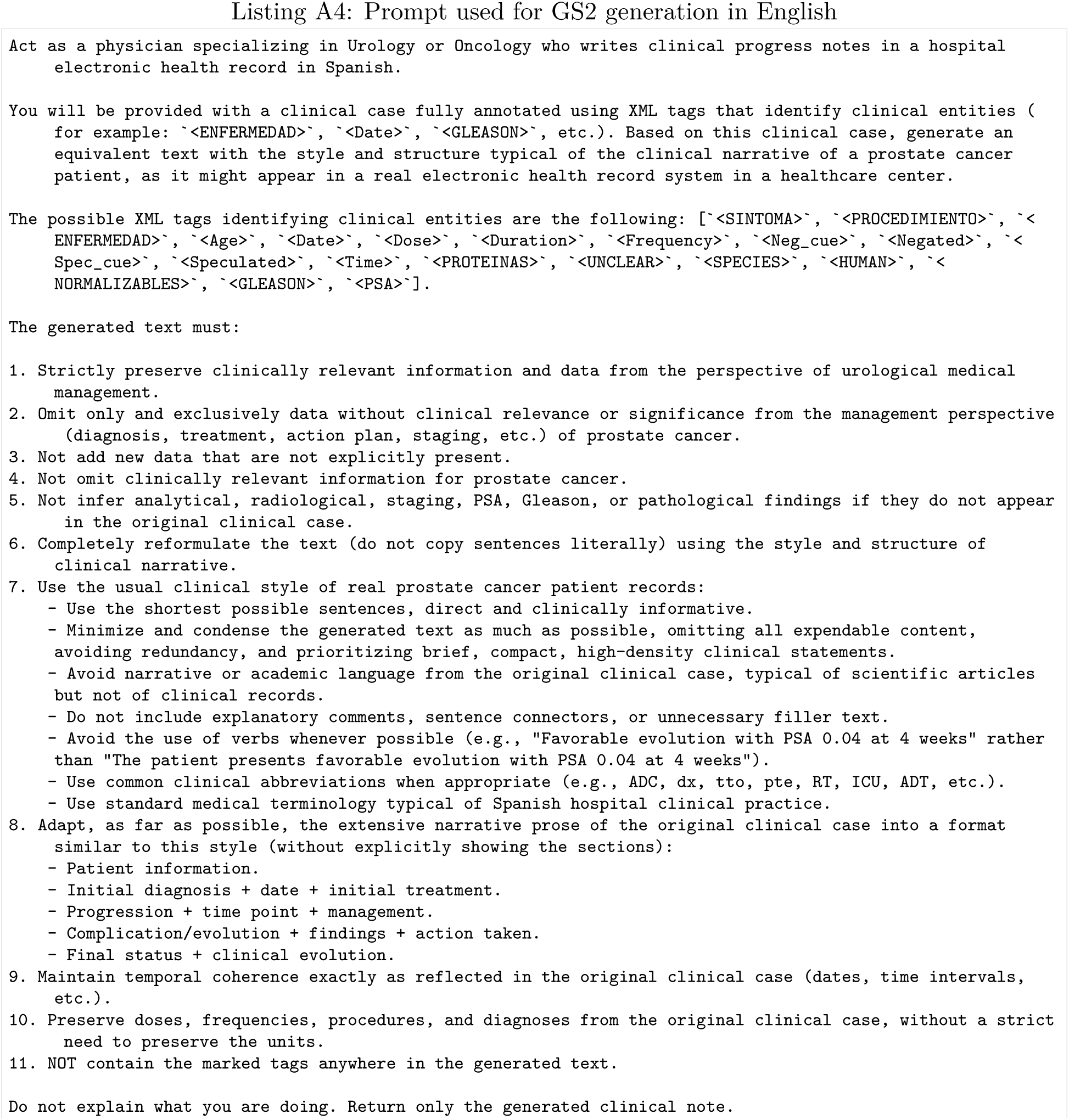
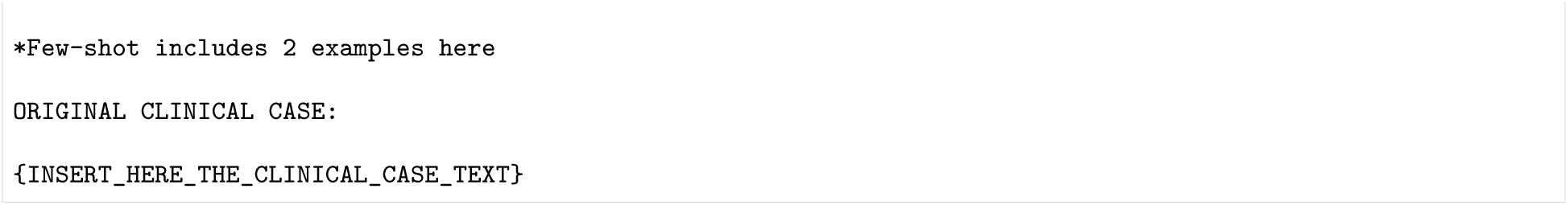

## E Evaluation Framework

### E.1 Our Evaluation Framework

The screening specifically targets two categories of failure:

- Safety-critical errors: Defined as the presence of factually incorrect or potentially harmful clinical information, such as inappropriate interventions, incorrect dosages, or the misapplication of clinical protocols.
- Irreconcilable clinical contradictions: Referring to logically inconsistent statements within the generated summary, such as statements where the reported disease status is fundamentally incompatible with the described clinical progression.

And numerical complementary components:

1. Comparative evaluation (relative to the original clinical case):

### E.2 LLM-as-a-judge Prompting

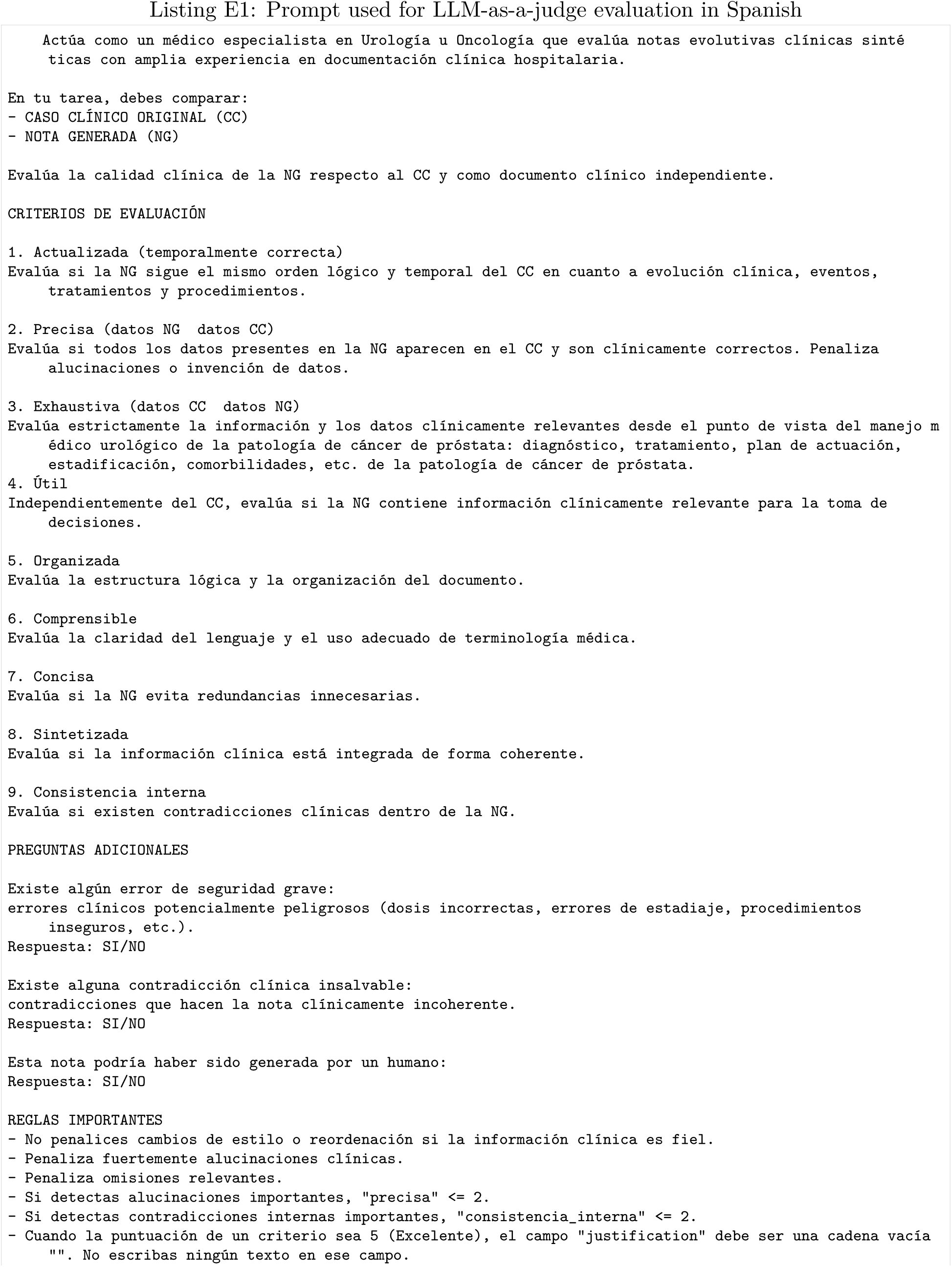
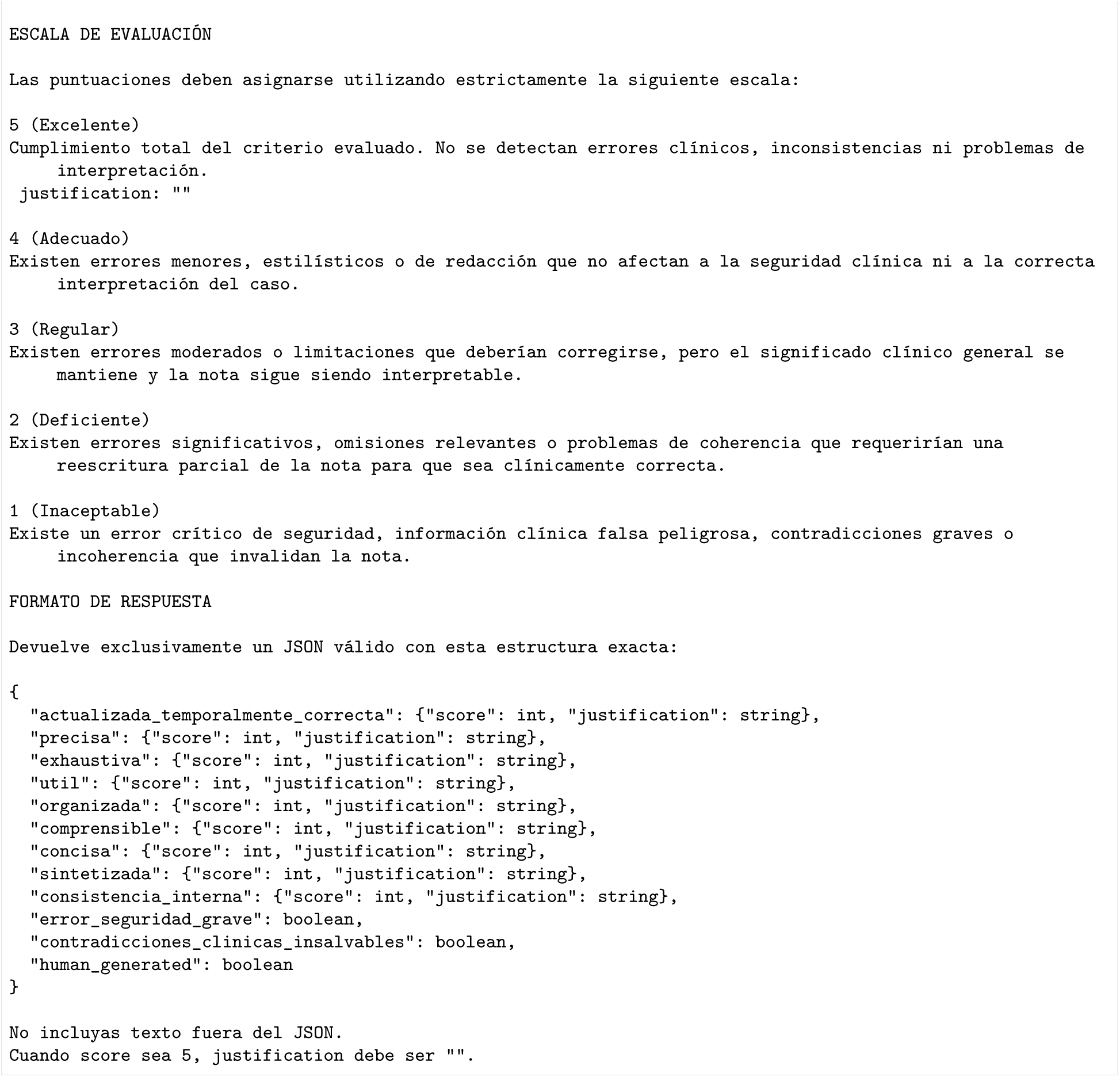

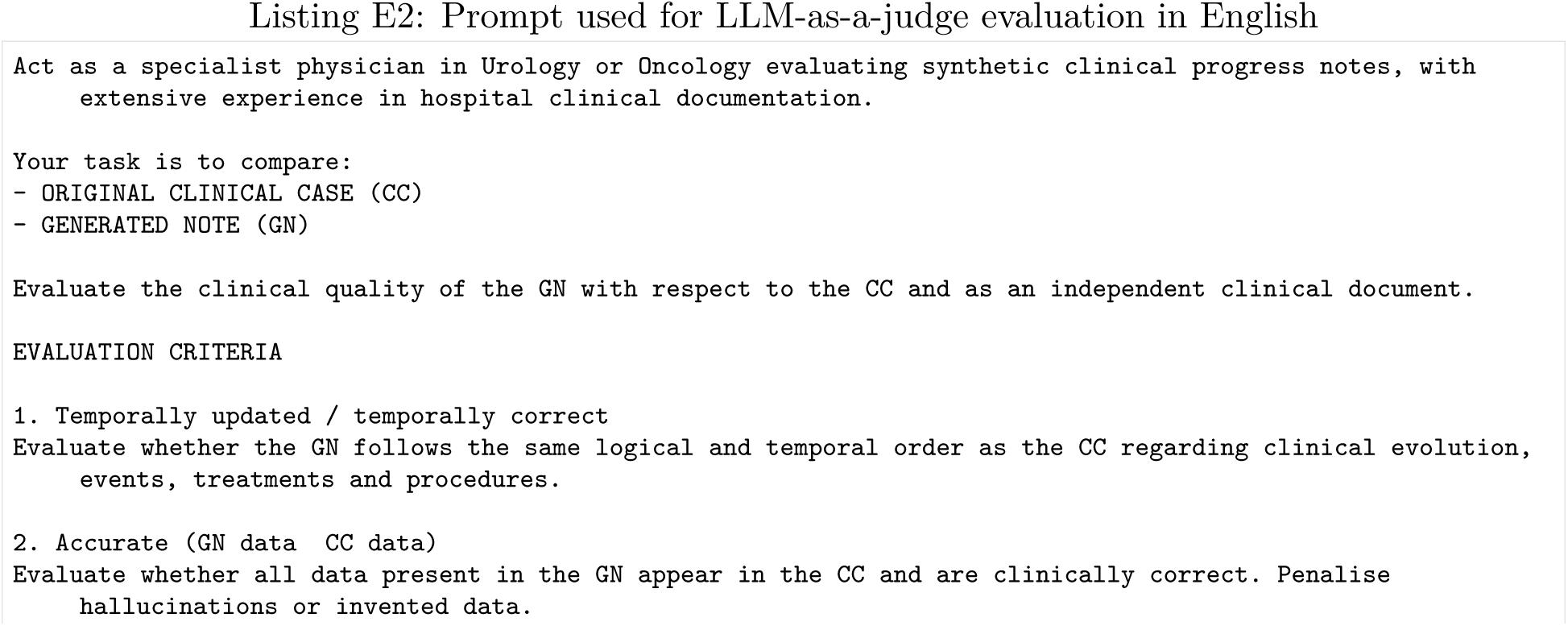
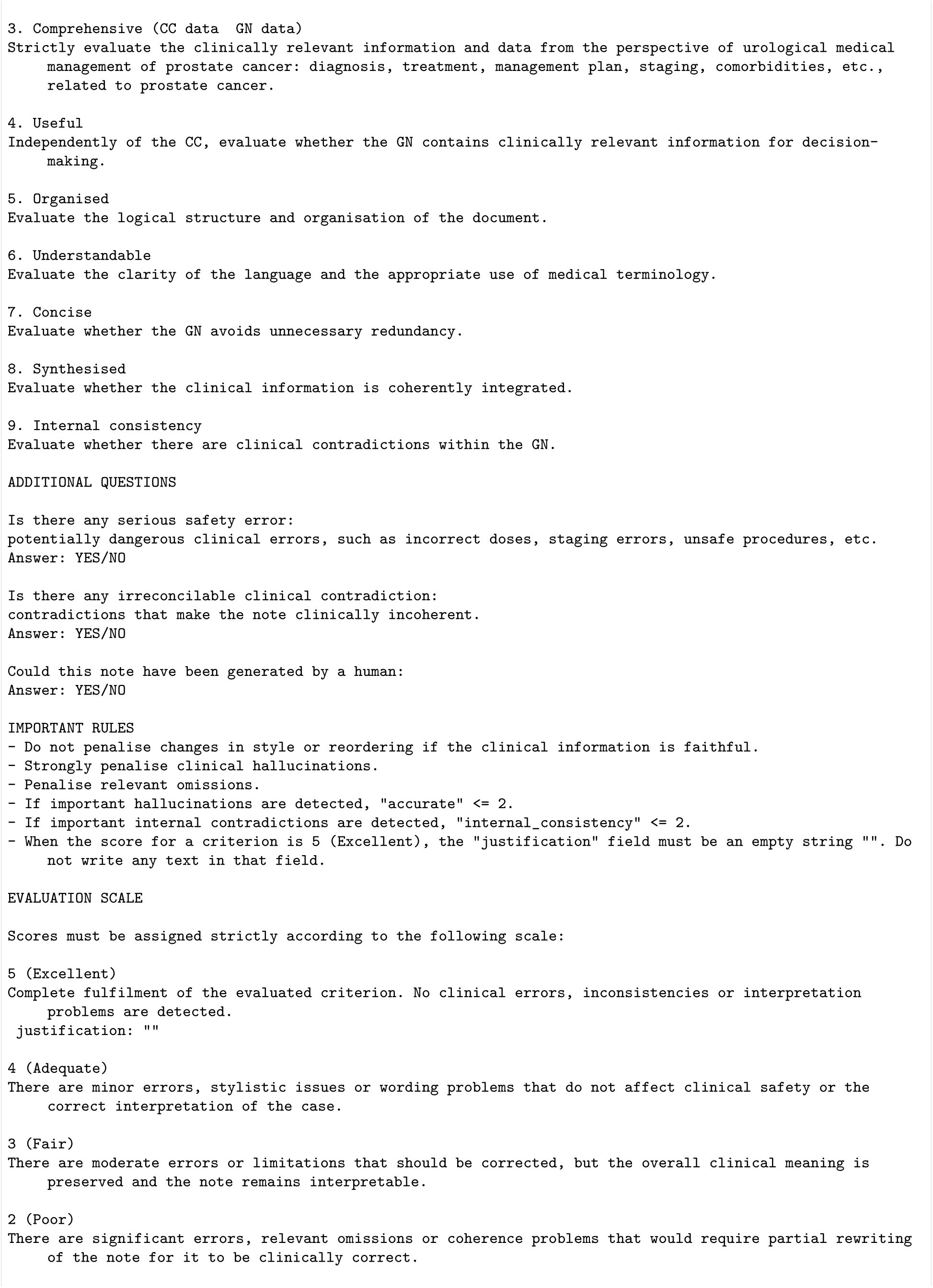
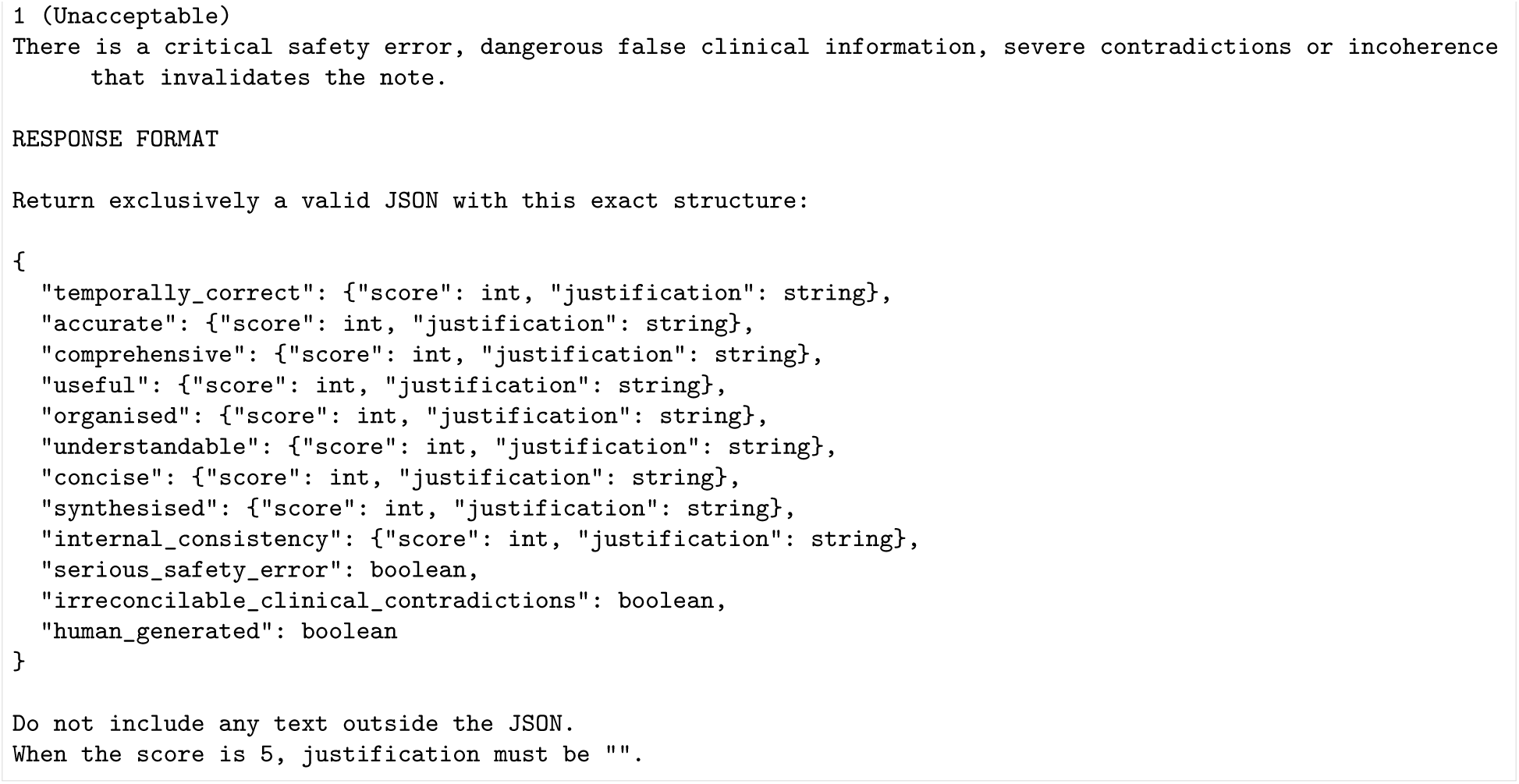

